# Neurofilaments as blood biomarkers at the preataxic and ataxic stage of spinocerebellar ataxia type 3: a cross-species analysis in humans and mice

**DOI:** 10.1101/19011882

**Authors:** Carlo Wilke, Eva Haas, Kathrin Reetz, Jennifer Faber, Hector Garcia-Moreno, Magda M. Santana, Bart van de Warrenburg, Holger Hengel, Manuela Lima, Alessandro Filla, Alexandra Durr, Bela Melegh, Marcella Masciullo, Jon Infante, Paola Giunti, Manuela Neumann, Jeroen de Vries, Luis Pereira de Almeida, Maria Rakowicz, Heike Jacobi, Rebecca Schüle, Stephan A. Kaeser, Jens Kuhle, Thomas Klockgether, Ludger Schöls, SCA3 neurofilament study group, European Integrated Project on Spinocerebellar Ataxias (EuroSCA/RiSCA), European Spinocerebellar Ataxia Type 3/Machado-Joseph Disease Initiative (ESMI), Christian Barro, Jeannette Hübener-Schmid, Matthis Synofzik

**Affiliations:** Hertie Institute for Clinical Brain Research (HIH) and Center of Neurology, University of Tübingen, Germany; German Center for Neurodegenerative Diseases (DZNE), University of Tübingen, Germany; Institute of Medical Genetics and Applied Genomics, University of Tübingen, Germany; Centre for Rare Diseases, University of Tübingen, Germany; Department of Neurology, RWTH Aachen University, Aachen, Germany; JARA-BRAIN Institute Molecular Neuroscience and Neuroimaging, Forschungszentrum Jülich, RWTH Aachen University, Aachen, Germany; Department of Neurology, University Hospital Bonn, Bonn, Germany; German Center for Neurodegenerative Diseases (DZNE), Bonn, Germany; Ataxia Centre, Department of Clinical and Movement Neurosciences, UCL Queen Square Institute of Neurology; National Hospital for Neurology and Neurosurgery, University College London Hospitals NHS Foundation Trust; Center for Neuroscience and Cell Biology, University of Coimbra, Coimbra, Portugal; Donders Institute for Brain, Cognition, and Behaviour, Department of Neurology, Radboud university medical center, Nijmegen, The Netherlands; Faculdade de Ciências e Tecnologia, Universidade dos Açores, Ponta Delgada, Portugal; Department of Neuroscience, and Reproductive and Odontostomatological Sciences, Federico II University Naples, Naples, Italy; Sorbonne Université, Institut du Cerveau et de la Moelle épinière (ICM), AP-HP, Inserm, CNRS, University Hospital Pitié-Salpêtrière, Paris, France; Department of Medical Genetics, and Szentagothai Research Center, University of Pécs Medical School, Pécs, Hungary; Spinal Rehabilitation Lab (SPIRE), IRCCS Fondazione Santa Lucia, Via Ardeatina 306, 00179, Rome, Italy; Service of Neurology, University Hospital Marqués de Valdecilla (IDIVAL), University of Cantabria (UC) and Centro de Investigación Biomédica en Red de Enfermedades Neurodegenerativas (CIBERNED), Santander, Spain; Department of Neuropathology, University of Tübingen, Germany; Department of Neurology, University Medical Centre Groningen, University of Groningen, Groningen, the Netherlands; First Department of Neurology, Institute of Psychiatry and Neurology, Warsaw, Poland; Department of Neurology, University Hospital of Heidelberg, Heidelberg; Neurology, Departments of Medicine, Biomedicine and Clinical Research, University Hospital Basel, University of Basel, Basel, Switzerland

**Keywords:** spinocerebellar ataxia type 3 (SCA3), presymptomatic stage, Neurofilament Light chain (NfL), phosphorylated Neurofilament Heavy chain (pNfH), Single molecule array (Simoa) technique

## Abstract

Spinocerebellar ataxia type 3 (SCA3) is a devastating multisystemic neurodegenerative disease for which targeted molecular therapies are coming into reach (e.g. antisense oligonucleotides). To pave the way for upcoming translational trials, easily accessible biomarkers in SCA3 are needed, particularly for subjects at the preataxic stage and cross-validated also in animal models. We hypothesised that serum neurofilaments might serve as blood biomarkers of disease progression in both human SCA3 and mouse models, expecting increased concentrations already at the preataxic stage. Serum neurofilament light (NfL) and phosphorylated neurofilament heavy (pNfH) levels were determined by ultra-sensitive single molecule array (Simoa) in cross-sectional samples of ataxic and preataxic SCA3 subjects and controls in two independent cohorts (ESMI cohort = cohort #1: n=160, EuroSCA/RiSCA cohort = cohort #2: n=89). Serum NfL and pNfH were also assessed in a 304Q SCA3 knock-in mouse model across presymptomatic and symptomatic disease stages (n=147). Ataxic SCA3 subjects showed increased serum NfL (p<0.001) and pNfH (p<0.001) levels in cohort #1, with NfL levels already increased in preataxic subjects (p<0.001). All these results were replicated in cohort #2 (all p<0.001). Cross-sectional NfL levels correlated with clinical disease severity (Scale for the Assessment and Rating of Ataxia [SARA]; r=0.43, p<0.001) and with longitudinal disease progression (annual SARA score change, ϱ=0.42, p=0.012). CAG count and age were significant predictors of individual NfL concentrations (each p<0.001). NfL levels in preataxic subjects increased with proximity to individual expected onset of ataxia (p<0.001), with significant elevations already 7.5 years before onset. Serum NfL and pNfH increases in SCA3 subjects were paralleled by similar changes in SCA3 knock-in mice, here also already starting at the presymptomatic stage and close to the onset of ataxin-3 protein increase. Serum concentrations of neurofilaments, particularly NfL, might provide easily accessible biomarkers of disease severity in both ataxic and preataxic SCA3 subjects and mice prior to conversion. Neurofilaments thus entail potential applications as progression, onset/proximity and treatment-response markers in both human and murine SCA3 trials.

## Introduction

Spinocerebellar ataxia type 3 (SCA3), also known as Machado-Joseph disease, is the most common dominantly inherited form of degenerative ataxia, caused by an expanded CAG repeat in the *ATXN3* gene and marked by irreversible decline of motor function already in mid-life (Costa Mdo and Paulson, 2012; Rub *et al*., 2013). Advances in the understanding of the toxic gain-of-function mechanisms underlying SCA3 neurodegeneration have opened a window for targeted molecular therapies (Paulson *et al*., 2017; Ramani *et al*., 2017). Particularly, interventions with antisense oligonucleotides (ASOs) targeting mutated *ATXN3* show promising results in mitigating the molecular, pathological and behavioural disease-associated changes in a SCA3 mouse model (McLoughlin *et al*., 2018). ASO treatments might allow preventing the neurodegenerative process even before the occurrence of clinical symptoms (Finkel *et al*., 2017; Winter *et al*., 2019). However, to pave the way for upcoming trials of these promising therapies, easily accessible, objective and sensitive outcome parameters are urgently needed to track disease progression in both the preataxic and ataxic stage of SCA3 disease. Such parameters require validation in large human SCA3 cohorts with standardised phenotyping as well as in SCA3 mouse models, as mouse models allow comprehensive neuropathological validation and preclinical intervention trials during both the presymptomatic and symptomatic disease stage.

In this cross-species biomarker study, we propose serum concentrations of neurofilament light (NfL) and phosphorylated neurofilament heavy (pNfH) as easily accessible, objective and sensitive blood biomarkers of disease severity in SCA3. Neurofilaments (Nfs) are neuron-specific cytoskeletal proteins, rapidly released upon neuronal damage and, with novel ultra-sensitive single molecule array (Simoa) assays, quantifiable in peripheral blood (Wilke *et al*., 2016; Khalil *et al*., 2018). Our previous work in a mixed cohort of repeat-expansion spinocerebellar ataxias (SCAs) indicated that blood concentrations of NfL in multisystemic repeat-SCAs are increased at the ataxic disease stage (Wilke *et al*., 2018). Like NfL, phosphorylated neurofilament heavy (pNfH) might also allow capturing neuronal disintegration and particularly axonal damage in neurodegenerative disease, possibly capturing differential features of the neurodegenerative process (Khalil *et al*., 2018; Wilke *et al*., 2019).

We hypothesised that serum Nfs might serve as blood biomarkers of disease severity in both human SCA3 and mouse models, expecting increased concentrations at both the ataxic and preataxic stage, with increases in preataxic subjects occurring particularly in proximity to the onset of ataxia. We measured serum Nf concentrations in cross-sectional samples of ataxic and preataxic SCA3 subjects and controls in two independent multicentric cohorts, using two independent ultra-sensitive single molecule array (Simoa) approaches, and correlated Nf levels with measures of disease severity. We expected the blood Nf increases in human SCA3 to be paralleled by blood Nf increases in SCA3 animal models, also starting already in the presymptomatic stage. We therefore assessed plasma NfL and pNfH also in a SCA3 knock-in mouse model (Martier *et al*.) (Haas et al., in preparation) across presymptomatic and symptomatic disease stages, correlating Nf plasma concentrations with the temporal course of phenotypic and neuropathological disease features, including brain ataxin-3 protein levels.

## Materials and methods

### Human cohorts

Serum Nf concentrations were assessed in cross-sectional samples from two independent multicentric international SCA3 cohorts. Cohort #1 was recruited by the ESMI consortium (European Spinocerebellar Ataxia Type 3/Machado-Joseph Disease Initiative, recruitment period: 2016-2018), comprising 83 SCA3 subjects and 77 healthy controls. Cohort #2 comprised 41 SCA3 subjects and 48 healthy controls from the EuroSCA (European integrated project on spinocerebellar ataxias) and RiSCA (Prospective study of individuals at risk for spinocerebellar ataxia) cohorts (recruitment period: 2008-2016) (Jacobi *et al*., 2013; Jacobi *et al*., 2015). The recruitment of these studies included subjects with genetically confirmed SCA3 (*ATXN3* repeat length ≥53; for determination of CAG repeat length, see Supplement 1), their first-degree relatives (i.e. siblings and children), and unrelated neurologically healthy controls. Based on their score on the Scale for the Assessment and Rating of Ataxia (SARA) (Schmitz-Hubsch *et al*., 2006), SCA3 mutation carriers were classified as either ataxic (SARA score ≥3, cohort #1: 75 subjects, cohort #2: 27 subjects) or preataxic (SARA score <3, cohort #1: 8 subjects, cohort #2: 14 subjects). Controls comprised mutation-negative first-degree relatives of SCA3 carriers and unrelated healthy individuals, all without symptoms or signs of neurodegenerative disease. Sample size calculation based on a piloting study had indicated that 15 ataxic SCA3 subjects and 15 controls would suffice to detect significant differences of NfL serum levels between groups (assuming α=0.01, β=0.01, equal group size, use of two-tailed non-parametric test) (Wilke *et al*., 2018). However, we here included all available SCA3 subjects in each cohort to assess associations of Nf levels with clinical and genetic variables. For 35 SCA3 subjects in cohort #2, longitudinal SARA scores were available. Demographic, clinical and genetic characteristics of both cohorts are detailed in Table 1. All subjects provided written informed consent prior to participation according to the Declaration of Helsinki. The study was approved by the local ethics committees of the University of Tübingen and the other study sites.

**Table 1.** Demographic, clinical and genetic characteristics of the human SCA3 cohorts.

| **cohort #1**<br>**(ESMI)** | **ataxic SCA3** | **preataxic SCA3** | **controls** |
| --- | --- | --- | --- |
| sample size (female) | 75 (56.0%) | 8 (87.5%) | 77 (41.6%) |
| age [years] | 52.3 (45.7-61.0) | 33.7 (30.1-36.9) | 43.5 (31.4-58.9) |
| reported age of onset [years] | 40.0 (33.0-46.5) | NA | NA |
| disease duration based on reported age of onset [years] | 11.0 (7.2-15.1) | NA | NA |
| SARA score | 13.0 (10.0-21.5) | 1.0 (0.5-2.0) | NA |
| repeat count (long allele) | 70 (67-71) | 70 (67-71) | NA |

| **cohort #2**<br>**(EuroSCA/RiSCA)** | **ataxic SCA3** | **preataxic SCA3** | **controls** |
| --- | --- | --- | --- |
| sample size (female) | 27 (48.1%) | 14 (42.9%) | 48 (52.1%) |
| age [years] | 50.9 (46.3-55.0) | 36.7 (28.2-45.5) | 40.7 (31.2-50.0) |
| reported age of onset [years] | 40.0 (35.3-46.0) | NA | NA |
| disease duration based on reported age of onset [years] | 10.9 (6.1-16.9) | NA | NA |
| SARA score | 11.5 (6.5-15.0) | 1.0 (0.5-1.5) | NA |
| repeat count (long allele) | 69 (66-70) | 69 (65-70) | NA |

Data are reported as median and interquartile range. NA: not applicable.

### Human serum and neuropathology

Blood samples were centrifuged (4,000 g, 10 min, room temperature). Serum was frozen at −80 °C within 60 min after collection, shipped and analysed without any previous thaw-freeze cycle. Post-mortem brain and spinal cord tissue of the SCA3 subject ID27925 was assessed for degeneration and polyQ aggregates (Supplement 2).

### Mouse model, murine plasma and neuropathology

We used the 304Q SCA3 knock-in mouse model containing a 304 trinucleotide repeat expansion in the murine *ATXN3* homolog, which was generated on the background of C5BL/6N mice (Charles River) by zinc finger technology (Carbery *et al*., 2010), as described in detail elsewhere (Martier *et al*.) (Haas et al., in preparation) (phenotype onset: 8 months of age; for a brief description, see Supplement 1). The study comprised 147 animals (56 heterozygous, 37 homozygous, 54 wildtype) with balanced sex within the genotypic groups. Animals were dissected at 2, 6, 12 and 18 months of age, following CO2 introduction. Blood was obtained by heart puncture with heparinised syringes, collected in 1.3 ml lithium heparin tubes, and centrifuged (3,000 g, 20 min, 4°C) to obtain plasma. Samples were then aliquoted (70 µl) and stored at −80°C until use. Cerebellum and frontal lobe tissue were homogenized in Precellys CK14 tubes on the Precellys 24 homogenisator (VWR) at 10% (w/v) in homogenization buffer (50 mM Tris [pH 8.0], 150 mM NaCl, 5 mM EDTA, containing proteinase inhibitor (cOmplete Protease Inhibitor Cocktail, Roche)). Cerebellar samples were further treated with ultrasonic waves for better homogenisation. The homogenised brain tissue was centrifuged at 25,000 g, 4°C, for 60 min. The supernatant was aliquoted and diluted 1:10,000 in sample diluent. We quantified tissue levels of soluble mutant ataxin-3 by time-resolved Förster resonance energy transfer immunoassay (TR-FRET) (Nguyen *et al*., 2013) and aggregated mutant ataxin-3 by filter retardation assay (Weber *et al*., 2017; Weishäupl *et al*., 2018) (Supplement 1).

### Phenotypic assessment of mice

Given the strong abnormal weight phenotype of our SCA3 mouse model, indicating severe underlying disease processes already early in the disease course, weight was determined longitudinally in all animals every two weeks. Additionally, in an independent cohort of 304Q SCA3 mice (n=45), evolution of coordination and balance was assessed by quantitative gait analysis using the Catwalk 8.1 gait analysis system (Noldus) (Supplement 1).

The experiments were conducted in accordance with the veterinary office regulations of Baden-Wuerttemberg, Germany (Regierungspräsidium Tübingen: HG3/13, HG2/18), the German Animal Welfare Act and the guidelines of the Federation of European Laboratory Animal Science Association, based on European Union legislation (directive 2010/63/EU).

### Neurofilament quantification

Nf concentrations were measured in duplicates by ultra-sensitive single molecule array (Simoa) technique on the Simoa HD-1 Analyzer (Quanterix, Lexington, Massachusetts). In cohort #1, we used the NF-light Advantage kit for NfL (Kuhle *et al*., 2016) and the pNF-heavy Discovery kit for pNfH quantification (Wilke *et al*., 2019), according to the manufacturer’s instructions (Quanterix). In cohort #2, we quantified Nf concentrations using an independent homebrew NfL-pNfH duplex assay (Supplement 1). Given the minute volumes which suffice for measuring both analytes by the NfL-pNfH duplex assay (70 µl per sample), we also used this duplex assay for quantifying Nf concentrations in murine plasma and murine brain lysates from cerebellum and frontal lobe. All samples were measured in duplicates (dilution for serum/plasma: 1:4, for lysates: 1:2000). Technicians were blinded to the genotypic and phenotypic status of the samples. Validation with 47 independent serum samples (13 SCA3 subjects, 34 controls) confirmed excellent agreement between the Quanterix and the homebrew NfL assays (R=0.99). This allowed transformation of the NfL homebrew measurements to the scale of the Quanterix measurements by linear regression (Supplement 3 for assay characteristics and validation). For pNfH, analogous validation also showed good, yet lower agreement between the Quanterix and the homebrew pNfH assays (R=0.88).

### Statistical analysis

*Group effects*. We used non-parametric procedures to analyse group effects on Nf levels (Mann Whitney U tests, two-sided, Bonferroni corrected for multiple comparisons). To correct the group effects for age-dependent increases of Nf levels (log-transformed) within each cohort, we used linear models with the factors group, age and their interaction. The optimal cut-offs for differentiating ataxic SCA3 subjects from controls by their Nf levels were determined according to Youden’s procedure, with the cut-off allowing to benchmark Nf levels in preataxic SCA3 subjects.

#### Associations with disease severity and disease progression

In ataxic subjects, we assessed the association of Nf levels (log-transformed) and disease severity, as captured by the SARA score, with Pearson’s correlations (two-sided test). We used partial correlations with the covariates age and disease duration to adjust the association between Nfs and disease severity for potential confounders. Prospective longitudinal SARA scores were available for a subset of patients (n=35; all from cohort #2; number of longitudinal visits per subject: 2-4) allowing us to determine intraindividual disease progression as the annual change of SARA scores (determined by intraindividual linear regression coefficients, using all available SARA scores). As values for disease progression did not meet the assumption of normality, we assessed the association between intraindividual longitudinal disease progression and cross-sectional Nf concentrations by Spearman’s correlation (two-sided test).

#### Association of NfL levels with age and repeat length

We analysed the association of NfL levels with age and CAG repeat length in SCA3 carriers with a linear model, using the pooled NfL data of both cohorts after transforming NfL levels of cohort #2 (which were measured by homebrew Simoa) to the scale of the Quanterix Simoa used for cohort #1. Specifically, we modelled NfL levels (log-transformed) in all SCA3 carriers (n=123) with the predictors age and *ATXN3* CAG repeat length, their squares and all possible interactions, analogous to previous analyses in Huntington’s disease (Byrne *et al*., 2017). We centred age at 50 years (i.e. mean age of carriers) and CAG repeat length at 68 (i.e. mean CAG repeat length of carriers), as in analogous previous analyses (Byrne *et al*., 2017). We excluded one outlier (NfL > 700 pg/ml) to fulfil model assumptions.

#### Temporal dynamics of NfL in preataxic SCA3

We analysed NfL levels as a function of time to estimated onset of ataxia in preataxic SCA3 by linear regression. For each preataxic SCA3 subject, we individually calculated the time to the estimated onset of ataxia based on CAG repeat count and age, as established previously (Tezenas du Montcel *et al*., 2014). To determine the point of time at which NfL levels become significantly increased in the preataxic stage of SCA3, NfL levels of preataxic carriers needed to be related to the NfL levels of controls at the same age, as NfL levels physiologically increase with age in controls (Wilke *et al*., 2016). Hence, we calculated the z-score of each SCA3 subject in relation to the NfL distribution in controls at the same age (for a visualisation, see Fig. 4B). For this, the difference between the measured NfL level and the NfL level estimated for controls at the same age was standardised relative to the NfL distribution in controls at this age. NfL in controls was modelled by linear regression on the level of log-transformed data.

#### Murine data

We compared biomarker levels (NfL, pNfH, aggregated ataxin-3, soluble ataxin-3) and phenotypic data between heterozygous and wildtype animals within the same age group (i.e. animals sacrificed at 2, 6, 12 or >12 months), using independent t-tests (two-sided, Bonferroni corrected for multiple comparisons given that two analytes were measured from each sample). Analogously, data were compared between homozygous mutants and wildtype animals.

Throughout the manuscript, if the assumption of normality was violated, we used log-transformed data for the statistical analysis after ensuring that the transformed data were normally distributed. We analysed the data with SPSS (IBM, Version 24). We reported the effect size r of the statistical tests wherever possible.

### Data availability

Data will be made available upon reasonable request and as patient consent allows. The authors confirm that the data supporting the findings of this study are available within the article and its Supplementary material.

## Results

### Serum NfL levels are increased at the ataxic stage of SCA3

In cohort #1 (Fig. 1A), serum concentrations of NfL were significantly higher in ataxic SCA3 subjects (34.8 pg/ml (28.3-47.0), median and IQR) than in controls (8.6 pg/ml (5.7-11.7)) (U=151, z=10.1, p<.001, r=0.82). In cohort #2 (Fig. 1B), NfL levels were also significantly higher in ataxic SCA3 subjects (85.5 pg/ml (70.2-100.2)) than in controls (19.4 pg/ml (15.1-25.4)) (U=16, z=6.98, p<.001, r=0.81). This confirmed the NfL increase in a second, independent cohort with an independent immunoassay. NfL levels differentiated between ataxic SCA3 subjects and controls with high accuracy (cohort #1: AUC=0.97 (0.95-1.00), p<.001, optimal cut-off: 20.0 pg/ml, 98.7% sensitivity, 92.2% specificity; cohort #2: AUC=0.99 (0.97-1.00), p<.001, optimal cut-off: 50.9 pg/ml, 92.6% sensitivity, 100% specificity). If corrected for age, the NfL increase in ataxic SCA3 subjects remained highly significant in both cohort #1 (F(1,147)=406.54, p<.001; based on a linear model with the factors group, age and their interaction, R^2^=0.82, Fig. 1C) and cohort #2 (F(1,70)=169.49, p<.001, R^2^=0.79, Fig. 1D).

**Fig. 1.**
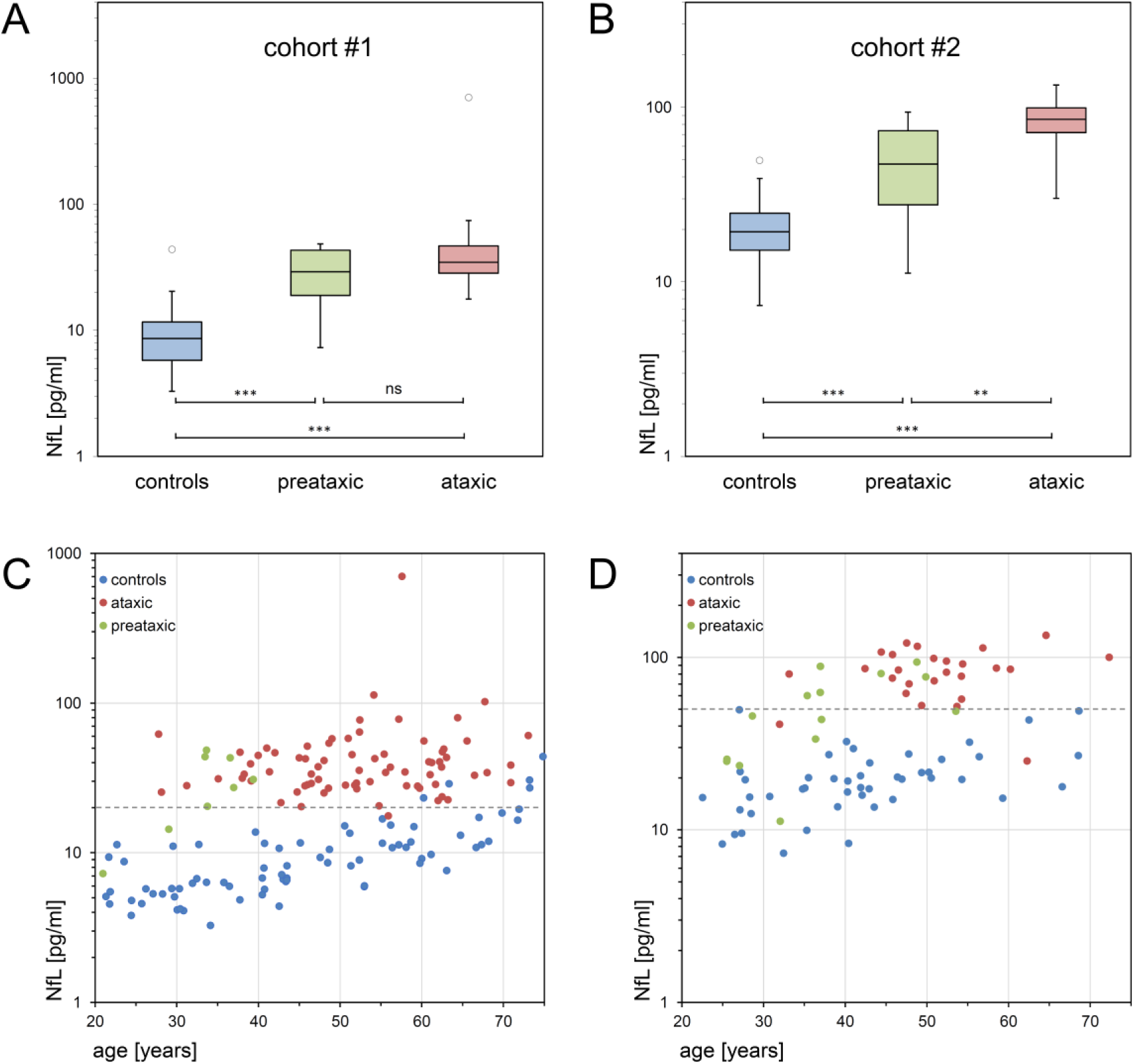
Serum NfL concentrations in the preataxic and ataxic stage of SCA3. Serum NfL concentrations of preataxic (green) and ataxic (red) SCA3 subjects and controls (blue) were measured in two independent cohorts, each with a different Simoa approach: cohort #1, recruited by the ESMI consortium (A, C), and cohort #2, recruited by the EuroSCA/RiSCA consortium (B, D). Boxes show the ranges between lower and upper quartiles, the central bands show the medians, and the whiskers show data within 1.5.IQR of the median, with dots representing outliers. Groups were compared with Mann Whitney U tests (*** p<.001, ** p<.01, ns p≥.05, Bonferroni-corrected). In the scatter plots, the individual NfL values were plotted as a function of subjects’ age. The dashed grey lines visualise the optimal cut-offs for differentiating ataxic SCA3 subjects from controls in each cohort (cohort #1: 20.0 pg/ml, 98.7% sensitivity, 92.2% specificity; cohort #2: 50.9 pg/ml, 92.6% sensitivity, 100% specificity; cut-offs were derived by maximising Youden’s index irrespective of age). Note the logarithmic scale of the y-axes.

### Serum NfL levels are increased at the preataxic stage of SCA3

In cohort #1 (Fig. 1A), NfL levels of preataxic SCA3 subjects (29.1 pg/ml (15.9-43.7)) were significantly higher than in controls (U=72, z=3.55, p<.001, r=0.39) and did not differ significantly from those of ataxic SCA3 subjects (U=204, z=1.48, p=.143, r=0.16, Bonferroni-corrected for multiple comparisons, respectively). In cohort #2 (Fig. 1B), NfL levels of preataxic SCA3 subjects (47.3 pg/ml (25.5-78.0)) were also significantly increased (U=88, z=4.18, p<.001, r=0.53), yet significantly lower than in ataxic SCA3 subjects (U=74, z=3.16, p=.001, r=0.49, Bonferroni-corrected). Within preataxic subjects, 75% (cohort #1) and 43% (cohort #2) of NfL levels were above the optimal cut-off separating ataxic subjects from controls. The NfL increase in preataxic SCA3 subjects remained highly significant if corrected for age, both in cohort #1 (F(1,81)=99.27, p<.001; based on a linear model with the factors group, age and their interaction, R^2^=0.68, Fig. 1C) and cohort #2 (F(1,58)=59.82, p<.001, R^2^=0.56, Fig. 1D).

### Serum NfL levels reflect disease severity

NfL levels of ataxic SCA3 subjects significantly correlated with disease severity (r=0.43, p<.001), as captured by the SARA score (Fig. 2A). The association between NfL and disease severity remained highly significant if corrected for age (r=0.41, p<.001), disease duration (r=0.41, p<.001) and CAG repeat length (r=0.47, p<.001) as possible confounders with partial correlations.

**Fig. 2.**
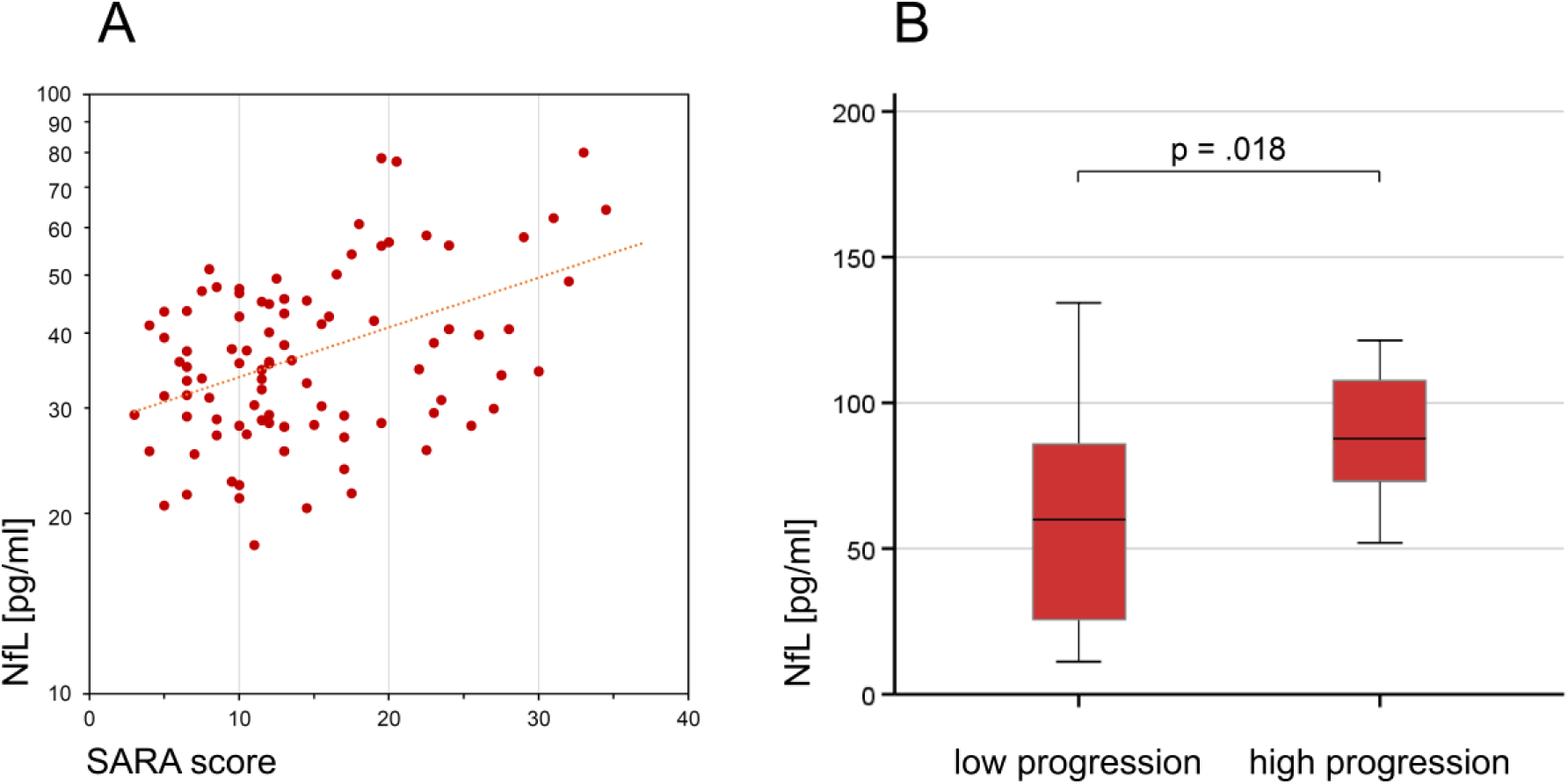
Associations of serum NfL levels with disease severity and progression. Serum NfL levels of ataxic SCA3 subjects significantly correlated with disease severity (A), as quantified by the Scale for the Rating and Assessment of Ataxia (SARA) score (r=0.43, p<.001, Pearson’s correlation). The cross-sectional NfL levels also reflected longitudinal disease progression, as quantified by the annual SARA score change (available for 35 subjects) (B). Subjects with high disease progression (annual SARA score increase ≥0.71 points/year, n=18, median split) had significantly higher serum levels of NfL than subjects with low disease progression (n=17) (p=.018, r=0.40, Mann Whitney U test).

### Serum NfL levels may reflect longitudinal disease progression

In a subset of ataxic SCA3 subjects (n=35, all from cohort #2), prospective longitudinal SARA scores were available to estimate intraindividual disease progression, as quantified by the annual change of the SARA score. Subjects’ cross-sectional NfL levels significantly correlated with the annual change of the SARA score (ϱ=0.42, p=.012), suggesting that serum NfL may also reflect longitudinal disease progression. Accordingly, subjects with high disease progression (annual SARA score increase ≥0.71 points/year, n=18, subset defined by median split) had significantly higher serum levels of NfL than subjects with low disease progression (n=17) (p=.018, r=0.40) (Fig. 2B).

### Association of NfL levels with repeat length and age

We analysed the association of NfL levels with age and CAG repeat length in SCA3 mutation carriers with a linear model, using the pooled data of both preataxic and ataxic subjects. The highly significant predictors of the NfL level were age (F(1,113)=40.54, p<.001), its square (F(1,113)=7.91, p=.006), and repeat length (F(1,113)=22.01, p<.001) (total explained variance: R^2^=0.37) (Fig. 3). The model demonstrated that, for a given age, each increase in the CAG repeat count was associated with higher NfL levels. For a given CAG repeat count, the NfL level increased with age, with the steepness of the slope declining with increasing age. Thus, the NfL increase in SCA3 reached a plateau in older age. The sustained increase of NfL levels through the ataxic stage was reflected by the absence of correlation of NfL levels with disease duration (Supplement 4). In controls, the relation between log transformed NfL levels and age was linear, indicating that any analyses comparing NfL levels between carriers and controls would need to consider the physiological NfL age-related increase in controls.

**Fig. 3.**
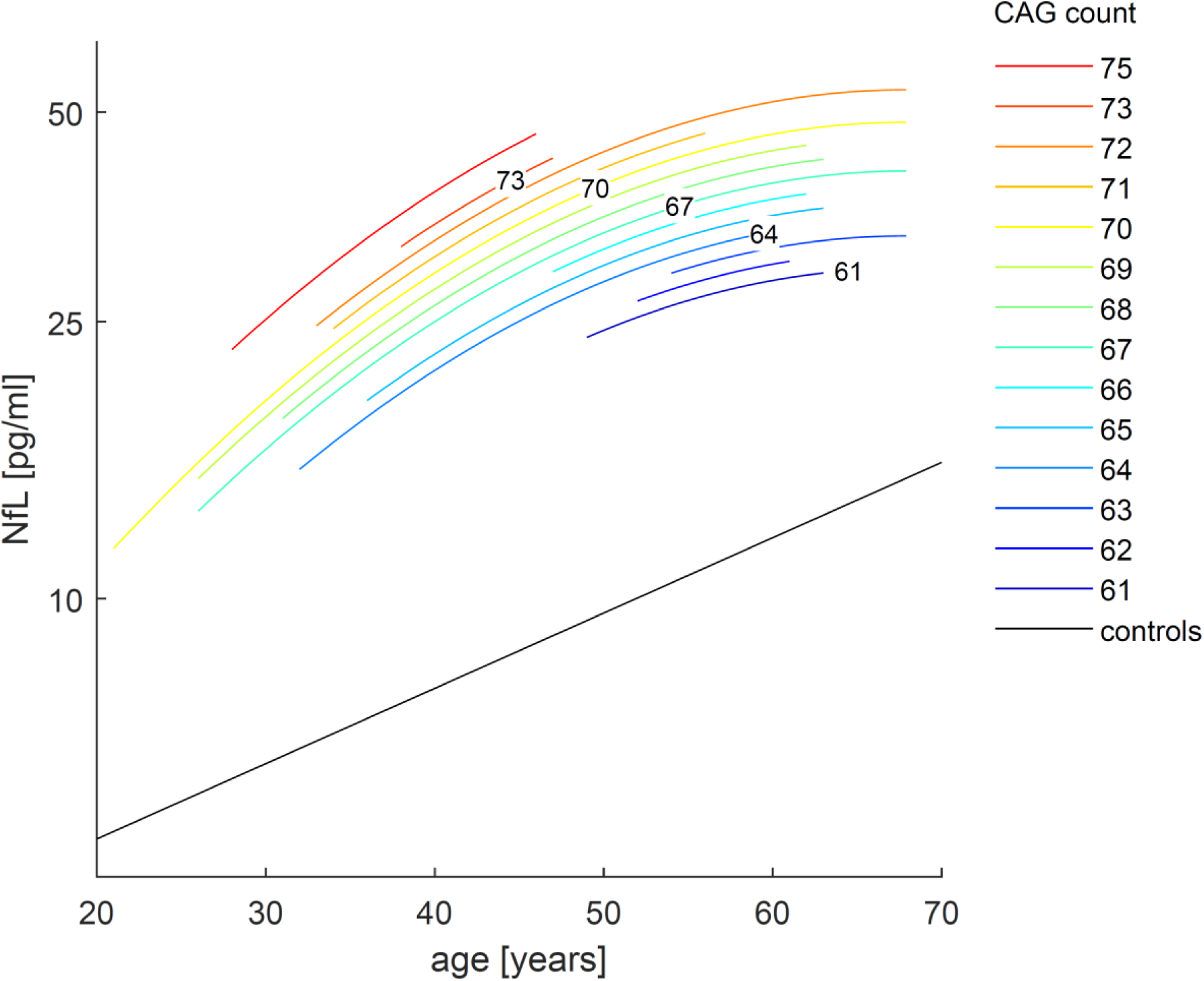
Association between NfL concentrations, age and CAG repeat length in SCA3. We modelled serum NfL levels (log-transformed) in SCA3 carriers (n=123) with the predictors age and *ATXN3* CAG repeat length, their squares and all possible interactions (for details, see main text). The highly significant predictors (age, its square, and repeat length, and the intercept, all p<.001, explained variance: R^2^=0.37) were used to generate the diagram. For a given age, each increase in CAG repeat count was associated with higher NfL concentrations. The steepness of the slopes declined with increasing age. In controls (black, n=125), the relation between NfL level (log-transformed) and age was linear.

### NfL levels increase with proximity to the estimated onset, with significant increases 7.5 years before onset of ataxia

NfL levels of preataxic subjects increased significantly with proximity to the individually predicted onset of ataxia, as revealed by a linear regression using the pooled data of both cohorts (F(1,21)=39.68, p<.001, R^2^=0.64; slope: 2.89 (1.94-3.85), p<.001) (Fig. 4A). To compare preataxic SCA3 subjects with controls at the same age, we expressed the measured NfL level of SCA3 subjects as NfL z-score in relation to the age-dependent NfL distribution in controls (Fig. 4B), and analysed the NfL z-score as a function of the time to the estimated onset of ataxia (Fig. 4C). The NfL z-score significantly increased with preataxic subjects approaching the expected onset of ataxia (F(1,21)=30.78, p<.001, R^2^=0.58; slope: 0.32 (0.20-0.44), p<.001) (Fig. 4C), without overlap of the 95% confidence interval of controls (i.e. if z-score > 1.96) already 7.5 years before the expected onset (Fig. 4C).

**Fig. 4.**
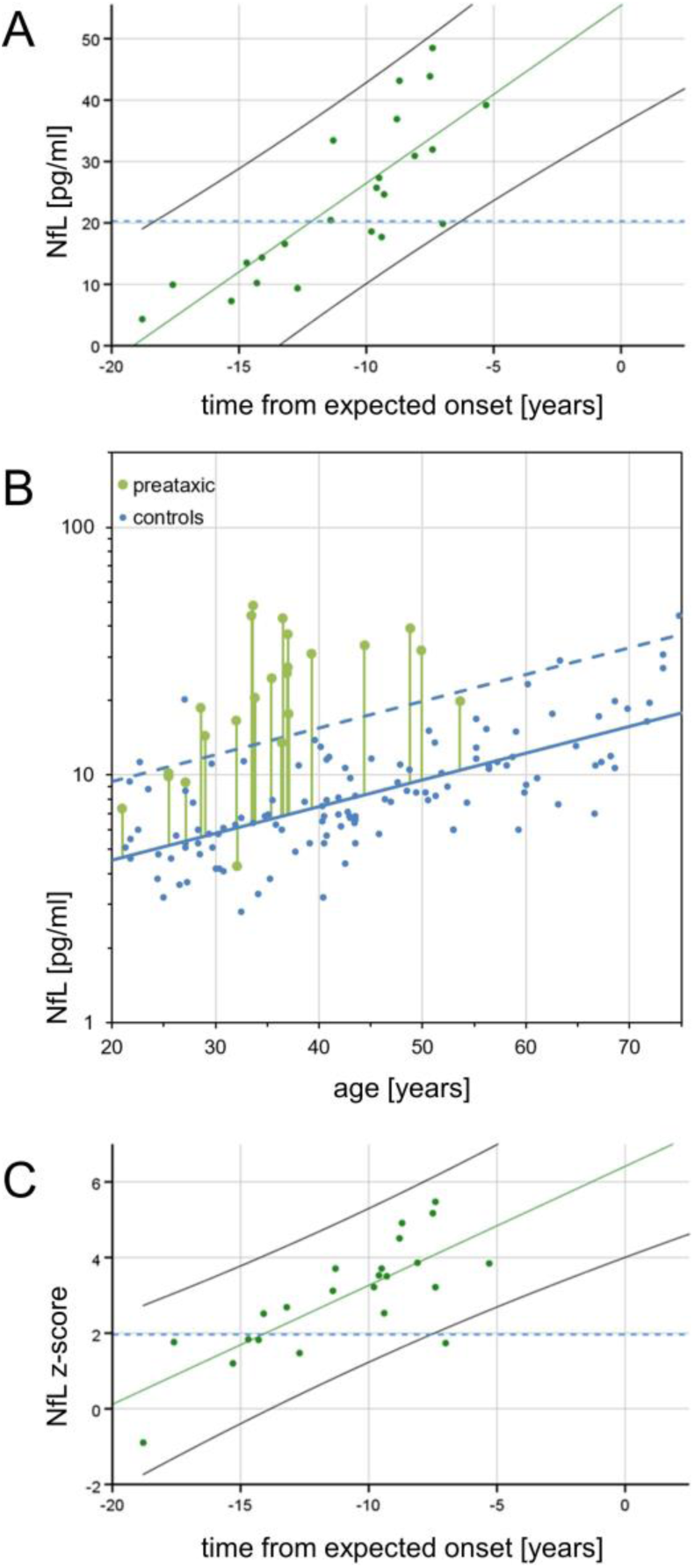
Temporal dynamics of serum NfL in preataxic SCA3. **(A)** Serum NfL levels in preataxic SCA3 subjects were plotted over the time from the individually estimated ataxia onset (A). NfL levels increased significantly with proximity to the estimated onset (F(1,21)=39.68, p<.001, R^2^=0.64). For benchmarking the levels of preataxic subjects, the blue dashed line visualises the optimal cut-off for differentiating ataxic SCA3 subjects from controls in the pooled cohort (20.3 pg/ml, 99.0% sensitivity, 95.2% specificity). **(B)** To determine the time at which NfL levels become significantly increased in the preataxic stage of SCA3, NfL levels of preataxic carriers need to be related to NfL levels of controls *at the same age*, as NfL levels physiologically increase with age. For each preataxic subject, the difference between the measured NfL value (green dot) and the NfL value predicted for control subjects of the same age (solid blue line) was visualised by the length of the vertical green line. Standardisation of this difference relative to the NfL distribution in controls (95% CI of the data: dashed blue line) yielded the individual NfL z-score which was plotted over each subject’s estimated time to onset in the next panel. **(C)** In preataxic SCA3 subjects, NfL z-scores increased significantly (F(1,21)=30.78, p<.001, R^2^=0.58) with subjects approaching the expected age of onset. NfL levels of preataxic subjects were significantly increased compared to controls (i.e. z-score > 1.96) 7.5 years before the expected onset, indicated by the non-overlapping 95% confidence intervals of SCA3 subjects (black solid line) and controls (blue dashed line).

### Using serum NfL to predict time to estimated onset of ataxia

The estimated time to onset might be predicted from individual NfL measurements (for z-scores in the range of 1 to 5) using the regression depicted in Fig. 4C. Moreover, the NfL z-score might allow delineating a preconversion stage, i.e. stratifying individual preataxic carriers close to expected symptom onset. The NfL z-score differentiated subjects at the late preataxic stage (i.e. carriers within 10 years of expected onset) from subjects at the early preataxic stage (i.e. more than 10 years before the expected onset) with high accuracy (AUC=0.89 (0.76-1.00) (95% CI), p=.002). Specifically, a cut-off for the NfL z-score at 3.2 differentiated early and late preataxic subjects with 85% sensitivity and 90% specificity.

### Increased phosphorylated neurofilament heavy (pNfH) levels in the ataxic disease stage of SCA3

In cohort #1 (Fig. 5A), SCA3 subjects at the ataxic stage had significantly higher serum pNfH concentrations (110.2 pg/ml (42.9-277.9)) than controls (22.8 pg/ml (13.9-59.1)) (U=1064, z=6.72, p<.001, r=0.55, Bonferroni-corrected), while pNfH concentrations in preataxic SCA3 subjects (33.9 pg/ml (18.0-184.8)) were not significantly increased compared to controls (U=201, z=1.61, p=.109, r=0.17). Likewise, in cohort #2 (Fig. 5B), serum pNfH levels were significantly higher in ataxic SCA3 subjects (23.2 pg/ml (3.2-71.2)) than in controls (3.2 pg/ml (3.2-10.4)) (U=260, z=4.57, p<.001, r=0.53), while pNfH concentrations in preataxic SCA3 subjects (4.5 pg/ml (3.2-34.6)) were not significantly increased (U=250, z=1.63, p=.103, r=0.21). These findings from two independent cohorts and assays validate pNfH increases in the ataxic stage of SCA3. Such increases might not necessarily be observed at the preataxic stage of SCA3. However, in both cohorts, pNfH levels at the preataxic stage were not significantly lower than in the ataxic stage of SCA3. While levels of NfL and pNfH were moderately correlated in controls (cohort #1: ϱ=0.28, p=.013, cohort #2: ϱ=0.39, p=.006), this correlation was only partially maintained in SCA3 mutation carriers (cohort #1, ϱ=0.13, p=.240, cohort #2: ϱ=0.33, p=.034), suggesting that pNfH might reflect a partly different feature/process in SCA3 than NfL.

**Fig. 5.**
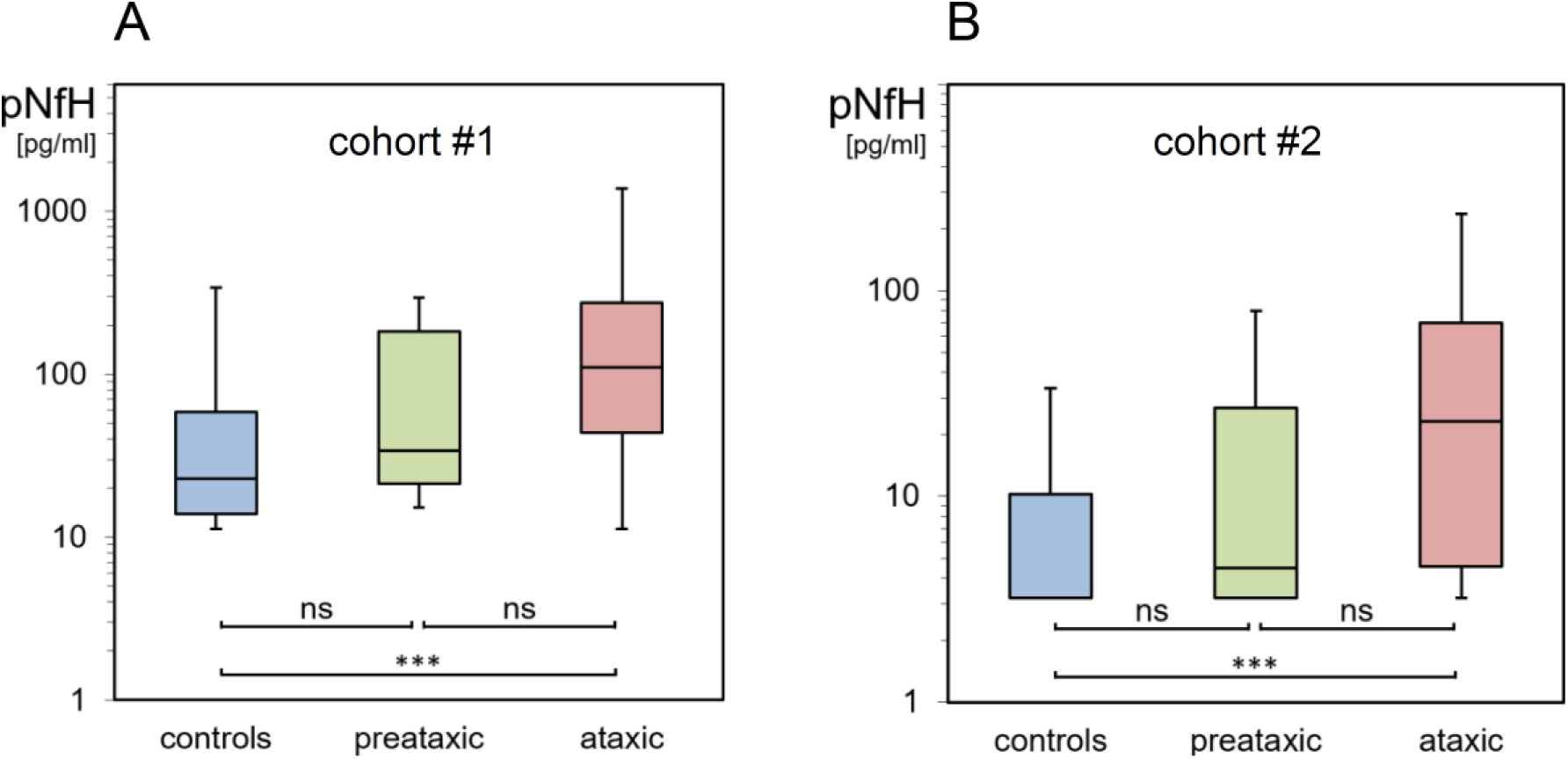
Serum pNfH concentrations in the preataxic and ataxic stage of SCA3. Serum pNfH concentrations of preataxic (green) and ataxic (red) SCA3 subjects and controls (blue) were measured in two independent cohorts with two different Simoa assays (A: cohort #1, B: cohort #2). Boxes show the ranges between lower and upper quartiles, the central bands show the medians, and the whiskers show data within 1.5.IQR of the median. Groups were compared with Mann Whitney U tests (*** p<.001, ns p≥.05, Bonferroni-corrected). Note the logarithmic scale of the y-axes.

### Human neuropathology

Neuropathologic examination of SCA3 subject ID27925 (cohort #2) with a NfL level of 116 pg/ml (93. percentile within the ataxic subjects of cohort #2) and a pNfH level of 23.2 pg/ml (52. percentile) at age 48.5 years (i.e. 12 months before death) did not show major degeneration of corticospinal tract, primary motor cortex and Purkinje cell layer, but marked degeneration of spinocerebellar tract, dentate nucleus and brainstem (Supplement 2). Similarly, while only rare polyQ pathology was found in the primary motor cortex and Purkinje cell layer, frequent polyQ pathology was present in the dentate nucleus, brainstem, basal ganglia and anterior horn. These findings, which correspond with and further corroborate previous SCA3 neuropathology case series (Paulson, 2012; Paulson *et al*., 2017; Koeppen, 2018), preliminarily indicate that not the corticospinal tract, motor cortex and cerebellar cortex, but rather affection of the spinocerebellar tract and brainstem might underlie the Nf increase in SCA3.

### Plasma neurofilament levels in a SCA3 mouse model are increased in proximity to phenotypic onset

To elucidate the temporal cascade of Nf concentrations in SCA3, we analysed blood concentrations of Nfs in a 304Q SCA3 knock-in mouse model with a phenotypic onset at around 8 months of age, sampling animals at age 2 months (= early presymptomatic), 6 months (= late presymptomatic), 12 months (= early symptomatic) and >18 months (= late symptomatic disease stage). While plasma concentrations of NfL were not yet increased at 2 months, they significantly increased already at age 6 months (p<.001) and 12 months (p<.001, Bonferroni-corrected) in heterozygous mutants compared to wildtype animals, with this effect levelling out in late symptomatic mice at age >18 months (Fig. 6A). Analogously, plasma concentrations of pNfH were significantly increased at 6 months (p<.01) and 12 months (p<.001) in heterozygous mutants compared to wildtype animals, but not at 2 months and >18 months, supporting an increase of Nf blood concentrations in proximity to the phenotypic onset (Fig. 6B). Exploratory analysis of Nf plasma levels in homozygous mutants confirmed the temporal cascade observed in heterozygous animals (Supplement 5). Tissue concentrations of NfL and pNfH in the cerebellum and frontal lobe did not differ between mutant and wildtype animals at the presymptomatic and symptomatic disease stage, preliminarily suggesting that the differences observed in the blood concentrations might relate not to changes of the amount of Nfs within the brain tissue, but rather to the *release* of Nfs from the brain tissue with neurodegenerative decay (Supplement 6). Brain Nf concentrations were hereby higher in the cerebellum than in the frontal lobe, suggesting higher share of axonal tissue in the cerebellum, and thus adding further support that *axonal* damage markers – like Nfs – might be promising markers for degenerative cerebellar disease.

**Fig. 6.**
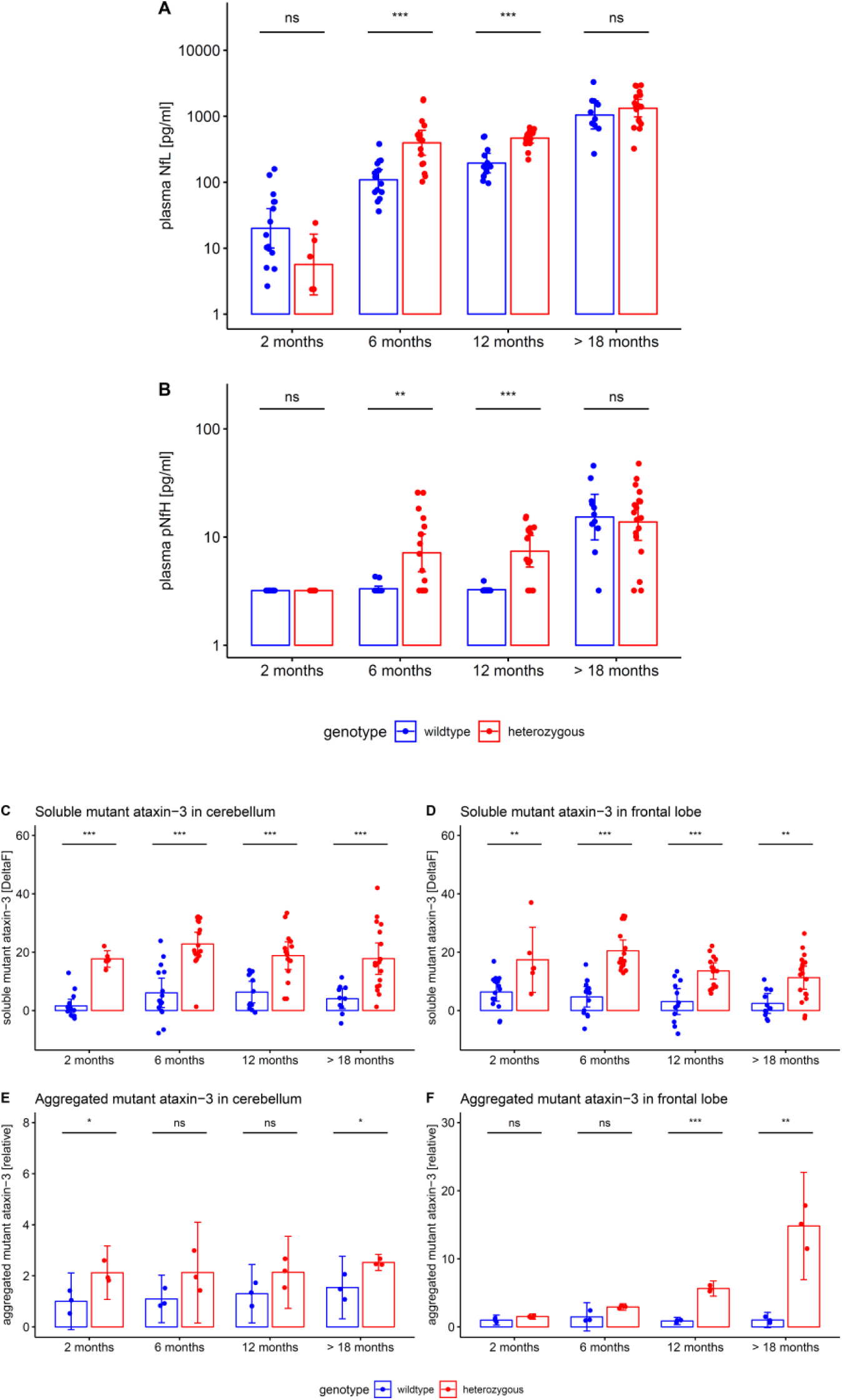
Neurofilament plasma levels and brain tissue levels of soluble and aggregated ataxin-3 in the 304Q SCA3 mouse model. Plasma concentrations of NfL (A) and pNfH (B) (log-transformed, mean and 95% CI) were measured in the 304Q knock-in SCA3 mouse model. Heterozygous animals become symptomatic at ≈ 8 months of age. Tissue levels of soluble and aggregated mutant ataxin-3 (mean and 95% CI) were measured in cerebellum (C, E) and frontal lobe (D, F). Heterozygous and wildtype animals were compared by unpaired t tests, adjusted for unequal variances (*** p<.001, ** p<.01, * p<.05, ns p≥.05, Bonferroni-corrected). Exploratory analyses in homozygous animals confirmed the ataxin-3 increases observed in heterozygous animals (Supplement 9).

Both NfL and pNfH increases in SCA3 relative to wildtype mice started earlier (age: 6 months) than the weight phenotype (age: 12 months) (Supplement 7) and the motor phenotype (age: 18 months) (Supplement 8), highlighting their value as proximity blood biomarkers for the preconversion stage also in mice. While Nf levels increased later than both aggregated and soluble cerebellar mutant ataxin-3 (age: 2 months) (Fig. 6), ataxin-3 levels did not change throughout the disease course and in particular not with proximity to conversion (Fig. 6). This indicates that, in contrast to Nf, ataxin-3 levels might serve less as preconversion markers, but possibly rather as target engagement markers. Taken together, our biomarkers and phenotype parameters allow mapping a preliminary multimodal chart of disease evolution in the 304Q SCA3 mouse model from the early preataxic to the late ataxic stages of SCA3 disease (Fig. 7).

**Fig. 7.**
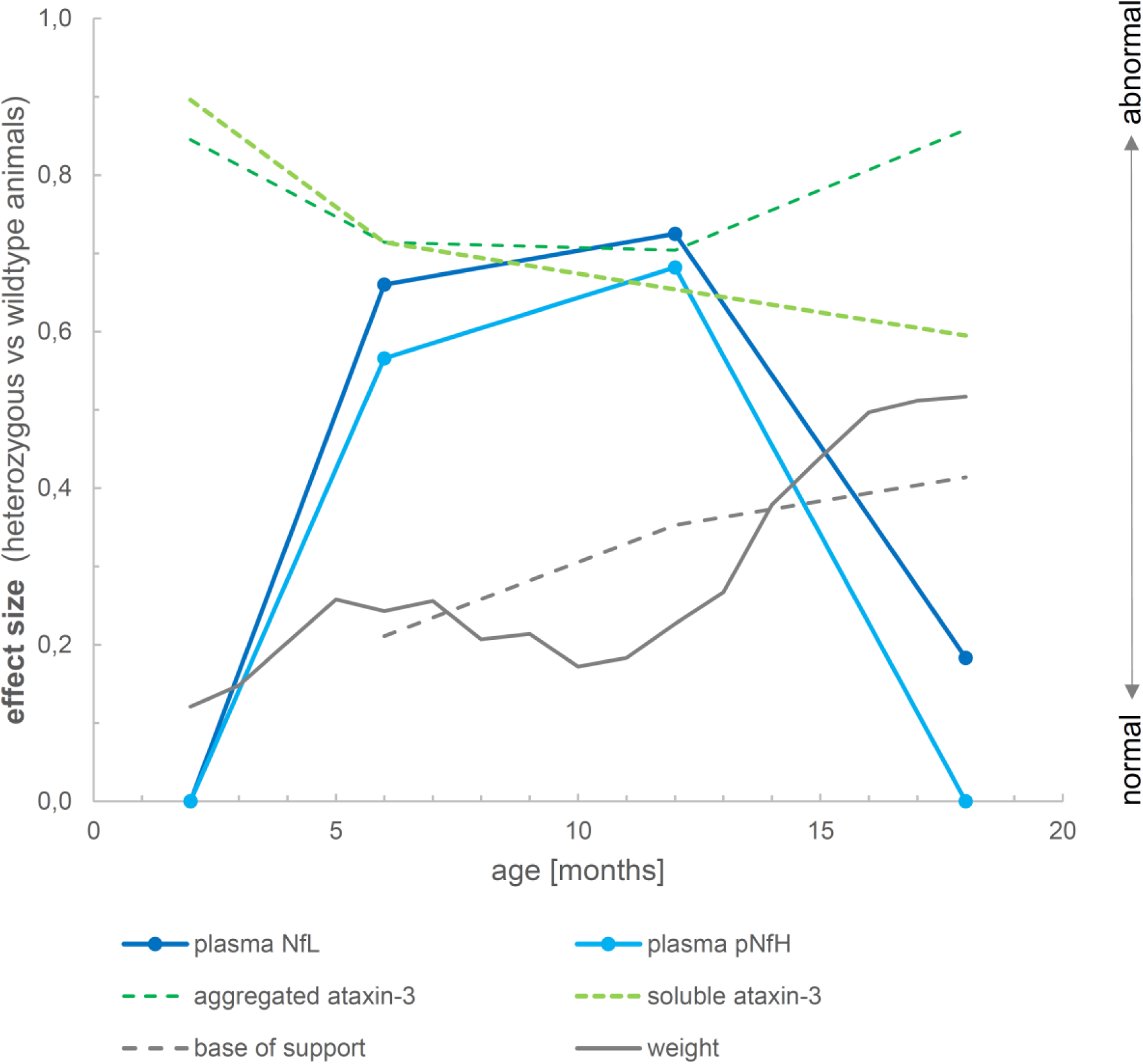
Biomarker cascade of NfL, pNfH, cerebellar aggregated and soluble ataxin-3 and phenotype in 304Q knock-in SCA3 mice. The figure shows the degree to which each variable differs between heterozygous SCA3 mice and wildtype controls for a given age. For each variable, we quantified the deviation of heterozygous mice from controls of the same age by calculating the effect size r of the group comparison (independent t-tests). An effect size of 0 indicates that the value in heterozygous mice is not different from controls, while an effect size of 1 would indicate strong abnormality. Note that the drop in the effect size of Nf levels from 12 months to 18 months thus does not imply a putative reduction of Nf levels, but rather the loss of the effect in mutants relative to controls, as controls show strong age-related Nf increases at 18 months of age. Negative effect size values were set to 0.

## Discussion

To pave the way for upcoming treatment trials, easily accessible surrogate biomarkers are highly warranted in SCA3 for both human and preclinical trials. In this comprehensive cross-species study – leveraging two independent large multicentric human cohorts and a knock-in mouse model, each covering both presymptomatic and symptomatic SCA3 disease stages and including both blood and neuropathology assessments – we demonstrate that (1) NfL levels are increased not only at the ataxic, but also in the preataxic stage of SCA3 in both humans and mice, and that (2) pNfH levels are increased at the ataxic stage of human SCA3 and at both the preataxic and ataxic stage of murine SCA3.

The observed increases of human NfL and pNfH serum levels in SCA3 seem to present a highly robust finding as they were each confirmed in two independent cohorts with two different Simoa assays. These Nf increases in SCA3 might not be primarily of help as a *diagnostic biomarker*, as Nfs are increased in many neurodegenerative and non-neurodegenerative conditions, including several multisystemic ataxias (Wilke *et al*., 2018). This is, however, not primarily needed for SCA3, as assessment of the *ATXN3* repeat expansion or the mutant protein meet this biomarker purpose. Rather, for preparing upcoming treatment trials, the biomarker value of Nfs might lie in their potential as *disease severity* and *stratification biomarkers*. We here demonstrate that NfL levels indeed correlate not only with cross-sectional disease severity, but also with the longitudinal annual change of the SARA score within individuals. These significant associations, though moderate in effect size, indicate that NfL blood levels might serve as easily accessible fluid biomarkers of disease severity in SCA3. Given that NfL levels distinguish fast versus slow disease progressors, they might also aid as stratification markers of SCA3 patients in upcoming treatment trials according to categories of disease progression. As NfL levels thus reflect disease severity and disease progression rates, but not clinical disease duration (Supplement 4), they seem to capture the functional stage and ongoing neuronal decay of SCA3 disease, rather than the mere calendrical estimate of duration of the disease. Increased NfL levels thereby might be related to spinocerebellar tract (and possibly brainstem) degeneration in SCA3, rather than e.g. corticospinal tract degeneration (as e.g. in ALS (Skillback *et al*., 2014; Menke *et al*., 2015)), as suggested by neuropathological findings. Our single-subject finding, however, requires validation in larger SCA3 cohorts with combined neuropathology and Nf assessment.

NfL levels appear to be strongly influenced, inter alia, by CAG repeat length and age, as both factors were found to present independent significant predictors of individual NfL levels. For a given age, each increase in the CAG repeat count was associated with higher NfL levels. This association of NfL levels and CAG repeat length suggests a dose-response relationship for SCA3 neurodegeneration and, together with analogous findings in Huntington’s disease (Byrne *et al*., 2017), more generally for polyglutamine disorders. For a given CAG repeat length, NfL levels increased with age, with the steepness of the slope decreasing with increasing age, i.e. the NfL increase in SCA3 reached a plateau in old age.

In both human cohorts, NfL levels were already increased at the preataxic stage of the disease, hereby the individual estimated time to onset might even be predicted from individual NfL levels. The NfL increase preceded the conversion to the ataxic stage by 7.5 years, with levels further increasing with proximity to the individually predicted onset of ataxia. NfL levels might thus allow to capture the onset of degenerative neuronal loss as well as the subclinical progression in the preataxic stage, possibly even on a single-subject level. They might moreover aid to stratify a window of opportunity in which future disease modifying therapies might be most effective. If future longitudinal assessments of converters confirm this NfL increase with proximity to ataxia onset (i.e. NfL as proximity/preconversion stratification biomarker), NfL might aid the selection of preataxic candidate subjects for therapeutic intervention, as invasive treatments (like ASOs) should likely be applied neither unnecessarily early nor too late in preataxic SCA3 subjects.

Serum levels of pNfH in both cohorts were increased at the ataxic stage of human SCA3, but – unlike NfL levels – not the preataxic stage. The differential timing of the pNfH increase might be, at least partially, related to higher biological variability of pNfH levels, given that assay characteristics of pNfH were similar to NfL. Given the higher sensitivity of NfL to capture changes already at the preataxic stage of human SCA3, our findings prioritise the use of NfL over pNfH in future SCA3 treatment trials. However, given their clear increase, both markers might still be further explored as combined fluid biomarkers in future longitudinal SCA3 trials, as they might show a differential response and dynamics to disease-modifying treatments or capture different features underlying multifaceted neurodegeneration, as suggested for other neurodegenerative diseases (Poesen and Van Damme, 2018; De Vivo *et al*., 2019; Winter *et al*., 2019).

Increases of NfL and pNfH blood levels in our two human SCA3 cohorts were paralleled by corresponding increases in our SCA3 mouse model, validating their value as neuronal damage biomarker in SCA3 *across species*. In mice, not only NfL, but also pNfH showed increases before the onset of the phenotype, namely at age 6 months. Like in human SCA3, these increases were sustained in SCA3 mice during the further disease course (i.e. at age 12 months), before the effects were then levelled out by the age-related increases of Nf in elderly wildtype mice (i.e. at age >18 months). Taking together our human and murine results, the temporal dynamics of Nf in SCA3 might hence be non-linear, with a boost of Nfs (in particular NfL) in proximity to phenotype conversion, and stabilisation of increased levels afterwards. Similar models of Nf dynamics have been proposed for other neurodegenerative diseases like amyotrophic lateral sclerosis (ALS) (Weydt *et al*., 2016) or frontotemporal dementia (FTD) (van der Ende *et al*., 2019). Hence, Nf levels (in particular NfL) might be particularly sensitive to neuronal decay in the *early* disease stage of SCA3, where biomarkers for tracking disease intensity would be most needed given the absence of change-sensitive clinical parameters.

The SCA3 mouse model also allowed us to relate the biomarker cascade of peripheral Nf levels to central ataxin-3 levels. Both soluble and aggregated mutant ataxin-3 increases in the cerebellum were detectable earlier than Nf increases, in line with the notion that they occur further upstream in SCA3 pathogenesis compared to the axonal damage reflected by Nf levels. Interestingly, in the frontal lobe aggregated mutant ataxin-3 seemed to increase later than Nf levels, indicating the Nf increase does not lag behind largely after ataxin-3 aggregation in some brain areas. In contrast to Nfs, however, ataxin-3 levels did not rise further during the late presymptomatic and early symptomatic stage in the SCA3 mice. This indicates that, while ataxin-3 might serve as a candidate biomarker for target engagement, it is of limited value as a disease severity or progression biomarker (in contrast to Nfs), at least in this SCA3 mouse model. Nfs and mutant ataxin-3 might thus overall serve as complementary biomarkers in preclinical SCA3 mouse treatment trials. Taken together, these comprehensive findings of our study largely extend recent preliminary findings from our and other labs (Wilke *et al*., 2018; Li *et al*., 2019) which suggested NfL increases in human SCA3, but were limited to non-multicentric, single-assay, human-only NfL studies, which also lacked the comparative assessment of NfL and pNfH and the biomarker cascade mapping.

Our study has several limitations. First, longitudinal assessment of Nf levels is warranted to confirm our cross-sectional Nf findings. The *intra*individual temporal dynamics of Nfs might be even more sensitive to early stages of degeneration and better suited to capture individual disease progression, as indicated by longitudinal Nf studies of genetic FTD and Alzheimer’s disease (van der Ende *et al*., 2019; Preische *et al*., 2019). Second, association of Nf levels with larger neuropathological and/or MRI datasets is warranted to determine the regions of the central nervous system which mainly drive the Nf increases. Yet, our study has identified the first set of fluid biomarker candidates for both human and preclinical treatment trials in SCA3.

## Data Availability

Data will be made available upon reasonable request and as patient consent allows.
Data will be shared with researchers who provide a methodologically sound proposal to achieve the aims in the approved proposal. Proposals should be directed to the corresponding author. To obtain data access, data requestors will need to sign a data access agreement.

## Acknowledgements

Biosamples were obtained from the Neuro-Biobank of the University of Tübingen, Germany, which is supported by the University of Tübingen, the Hertie Institute for Clinical Brain Research (HIH) and the German Center for Neurodegenerative Diseases (DZNE). CW was supported by the National Ataxia Foundation and the Wilhelm Vaillant Stiftung. CW, HH, BW, AD, TK, RS, LS and MS are members of the European Reference Network for Rare Neurological Diseases Project ID No 739510.

## Author contributions

CW: design and conceptualisation of the study, acquisition of data, analysis of the data, drafting and revision of the manuscript.

EH: design and conceptualisation of the study, conduction of animal experiments and acquisition of data, analysis of the data, revision of the manuscript.

KR: design and conceptualisation of the study, subject recruitment, acquisition of data, revision of the manuscript.

JF, HGM, MMS, BW, HH, ML, AF, AD, BM, MM, JI, PG, JV, LP, MR, HJ, LS: subject recruitment, acquisition of data, revision of the manuscript.

MN: acquisition of neuropathological data, analysis of the data, revision of the manuscript. RS, JK: acquisition of data, revision of the manuscript.

SK: design and conceptualisation of the study, revision of the manuscript.

TK: design and conceptualisation of the study, subject recruitment, acquisition of data, revision of the manuscript.

CB: design and conceptualisation of the study, acquisition of data, analysis of the data, revision of the manuscript.

JH: design and conceptualisation of the study, conduction of animal experiments and acquisition of data, analysis of the data, revision of the manuscript.

MS: design and conceptualisation of the study, acquisition of data, analysis of the data, drafting and revision of the manuscript.

**Appendix**

The SCA3 neurofilament study group includes Christian Deuschle, Elke Stransky, Kathrin Brockmann, JÖrg B. Schulz, Laszlo Baliko, Judith van Gaalen, Mafalda Raposo.

## Supplementary material

### Supplement 1. Supplementary methods

#### Genotyping of SCA3 subjects

The repeat length of the expanded and normal alleles was determined by PCR-based fragment length analysis from 100–250 ng genomic DNA (CEQ8000 capillary sequencer, Beckman Coulter). CAG repeat length was determined in a centralised manner for cohort #1.

#### Generation of the SCA3 knock-in mouse model

The SCA3 knock-in mouse model containing a 304 trinucleotide repeat expansion in the murine *ATXN3* homolog was generated on the background of C5BL/6N mice (Charles River) by zinc finger technology. In short, this method resulted in a double strand break within the murine (CAACAGCAG)2 region and the introduction of a specific expansion of the CAACAGCAG-region accomplished by homologous recombination using a donor vector with (CAACAGCAG)48 repeats, flanked by 800 bp up- and down-stream of *ATXN3*. More repeats than contained in the donor vector were introduced, resulting in 304 glutamines in the ataxin-3 protein. We used an interrupted CAG repeat (in the form of a (CAACAGCAG)n repeat) to ensure meiotic stability of the repeat length and to prevent repeat-associated non-AUG (RAN) translation, i.e. the model reflected polyglutamin pathology. We genotyped animals (n=147) by PCR, using DNA extracted from ear biopsies with the High Pure PCR Template preparation Kit (Roche). Animals were housed under standard conditions in groups (≤5 per cage) and maintained within a 12-hour light-dark cycle. Access to food and water was given ad libitum.

#### Quantification of soluble ataxin-3

Soluble mutant ataxin-3 measurements via time-resolved Förster resonance energy transfer immunoassay (TR-FRET) were performed using a combination of an anti-ataxin-3 antibody (clone 1H9, MAB5360, Merck) labelled with Tb (donor) fluorophore and an anti-polyQ antibody (clone 1C2, MAB1574, Merck) labelled with d2 (acceptor) fluorophore (labelling by Cisbio). Briefly, homogenized cerebellar and frontal lobe samples for Nf measurements were diluted in homogenisation buffer (50 mM Tris, 150 mM NaCl, 5 mM EDTA, cOmplete Protease Inhibitor Cocktail) to a final concentration of 1 µg/µl total protein amount. Next, 5 µl of diluted sample were incubated with 1 µl of the TR-FRET antibody mix (1 ng 1H9-Tb + 3 ng MW1-d2 in 50 mM NaH2PO4, 400 mM NaF, 0.1% BSA, 0.05% Tween-20) in a low-volume white ProxiPlate 384 TC Plus plate (PerkinElmer) at 4°C for 22 h. Signals were detected at 620 nm and 665 nm using an EnVision Multimode Plate Reader with a TRF-laser unit (PerkinElmer).

#### Quantification of aggregated ataxin-3

For filter retardation assay 12.5 µg of protein homogenate were diluted in Dulbecco’s phosphate buffered saline (DPBS) (Gibco) containing 2% sodium dodecyl sulphate (SDS) and 50 mM 1,4-dithiothreitol (DTT) and heat denatured for 5 min at 95°C. Samples were filtered through Amersham Protran Premium 0.45 µm nitrocellulose membranes (GE healthcare) by using a Minifold II Slot Blot System (Schleicher & Schuell). Before loading the samples, the membrane was equilibrated with DPBS containing 0.1% SDS and rinsed afterwards twice with DPBS. The membranes were blocked in tris-buffered saline (TBS) (1M Tris, 5M NaCl) containing 5% skimmed milk powder (Sigma-Aldrich) for 1 hour followed by primary antibody incubation (mouse-anti-Ataxin-3, 1:2500, clone 1H9, MAB5360, Merck Millipore) over night at 4°C. Secondary antibody (IRDye 800CW goat anti-mouse IgG (H+L) 1:1000, 926-32210, LICOR) incubated for 1.5 hours at room temperature. Detection and quantification of the fluorescent signal was performed using the ODYSSEY Server software version 4.1 (LI-COR Bioscience).

#### Motor phenotype assessment of the SCA3 mouse model

In an independent cohort of 304Q SCA3 mice (n=45), evolution of coordination and balance was assessed by quantitative gait analysis using the Catwalk 8.1 gait analysis system (Noldus). Animals were tested every six months starting with six months of age, 12-15 mice per genotype were analysed per time point. Each mouse had to complete five runs with a minimal run duration of 0.5 s and maximum of 10 s. The speed variation had to be below 60%. Run performance and footprint analysis were performed with Catwalk X software (Noldus).

#### Homebrew NfL-pNfH duplex Simoa assay

NfL and pNfH were measured by an in-house multiplex Simoa assay. For NfL, the monoclonal antibody (mAB) 47:3 was used as capture antibody and mAB 2:1 as detector antibody (UmanDiagnostics, Umeå, Sweden). pNfH was captured by mouse mAB anti-human-pNfH (Iron Horse Diagnostics, United States) and detected by chicken polyclonal anti-human pNfH antibodies (Iron Horse Diagnostics). Each capture antibody was separately coupled to dye encoded paramagnetic beads using a procedure previously described (Disanto *et al*., 2017). Bovine lyophilized NfL was obtained from UmanDiagnostics and purified pNfH from Ironhorse. Calibrators ranged for NfL from 0 to 2,000 pg/ml, for pNfH from 0 to 400 pg/ml (calibrator diluent: tris base saline (TBS); 0.1% Tween 20; 1% milk powder; 300 µg/mL Heteroblock (Omega Biologicals Inc., Bozeman, USA)). Batch prepared calibrators were stored at −80°C. Calibrators (neat) and samples (serum/plasma 1:4 dilution, lysates 1:2000 dilution; sample diluent: TBS, 0.1% Tween 20, 1% milk powder, 400 µg/mL Heteroblock (Omega Biologicals)) were measured in duplicates. Reagents were prepared as follows: NfL and pNfH beads were diluted to 1×104 beads/µl each and detector antibodies for NfL, pNfH were adjusted to 0.1 µg/ml and 0.05 µg/ml, respectively (beads and detectors diluent: TBS; 0.1% Tween 20; 1% milk powder; 300 µg/ml Heteroblock (Omega Biologicals)). Parallelism of the assay for serum, plasma and brain lysates were confirmed by serial dilution experiments in native samples.

### Supplement 2. Neuropathologic assessment of degeneration and polyQ pathology in a SCA3 subject with increased Nf levels

#### Methods

Post-mortem tissue of an ataxic SCA3 subject (ID27925), for whom also Nf levels were available, was provided by the brain bank affiliated with the University Hospital of Tübingen. Clinical assessment of subject ID27925 at the age of 48.5 years (ataxia onset: 34 years, disease duration: 15 years, death: 49.5 years, repeat length: 72) showed severe clinical affection (SARA score: 32) and above-average disease progression (2.2 SARA points/year; average progression in SCA3: 1.56 SARA points/year (Jacobi *et al*., 2015)). The subject showed serum NfL levels of 116 pg/ml (93. percentile within the ataxic subjects of cohort #2) and serum pNfH levels of 23.2 pg/ml (52. percentile). Histological examination was performed on 4 µm thick sections cut from formalin-fixed, paraffin-embedded tissue of frontal cortex, temporal cortex, primary motor cortex, putamen/pallidum, midbrain, pons, medulla, cerebellum and spinal cord. Sections were stained with haematoxylin and eosin (H&E), and Luxol fast blue–periodic acid–Schiff, or used for immunohistochemistry using the Ventana BenchMark XT automated staining system with the OptiView DAB detection kit (Ventana). Antibodies employed include monoclonal mouse anti-CD68 clone PG-M1 (Dako) and anti-polyQ clone 1C2 (Millipore). A semiquantitative grading system was used to score the severity of polyQ pathology (neuronal nuclear inclusions and granular cytoplasmic staining) as absent, mild (only few inclusions seen in the entire region examined), moderate (at least a few inclusions present in most microscopic fields) or severe (many inclusions in every microscopic field). Degeneration of brain regions and fibre tracts was assessed on H&E, myelin and CD68 stained sections and graded as absent, mild, moderate, severe based on the presence of spongiosis, neuronal loss, gliosis and/or myelin loss.

#### Results

Severity and distribution of degeneration and polyQ pathology are summarised in the table below (semiquantitative score: - absent, + mild, ++ moderate, +++ severe, NA: not applicable). The neuropathological findings indicate, in sum, that the spinocerebellar tract, brainstem and basal ganglia, but not the corticospinal tract, motor cortex or cerebellar cortex were mainly affected in this subject. These findings, which correspond with and confirm the findings of previous SCA3 neuropathology case series (Paulson, 2012; Paulson *et al*., 2017; Koeppen, 2018), might provide first preliminary indications of the central nervous system regions underlying the Nf increase in SCA3.

| CNS region | degeneration | polyQ pathology |
| --- | --- | --- |
| **cerebellum** |  |  |
| Purkinje cells | + | - |
| dentate nucleus | ++ | +++ |
| **brainstem** |  |  |
| pontine nuclei | ++ | +++ |
| substantia nigra pars compacta | ++ | ++ |
| oculomotor nuclei | ++ | ++ |
| inferior olive | + | +++ |
| XII nucleus | + | +++ |
| **cortex** |  |  |
| basal ganglia | + | +++ |
| primary motor cortex (layer V giant Betz cells) | + | + |
| frontal | - | + |
| temporal | - | + |
| **spinal cord** |  |  |
| anterior horn | ++ | +++ |
| **fibre tracts** |  |  |
| corticospinal tracts | + | NA |
| spinocerebellar tracts | ++ | NA |
| pontocerebellar fibres | ++ | NA |
| cerebellar peduncles | ++ | NA |

### Supplement 3. Analytical characteristics and validation of the neurofilament assays

In line with previous studies (Kuhle *et al*., 2016; Wilke *et al*., 2019), analytical sensitivity of each assay was defined as the analyte concentration of the calibrator with the lowest concentration fulfilling established acceptance criteria (i.e. coefficient of variation (CV) of duplicate determination ≤20% and accuracy within the range of 80-120%). Analytical sensitivity was corrected by multiplication with the dilution factor (i.e. multiplication with factor 4). Within-run precision and between-run precision were derived from four consecutive runs with samples of different analyte concentrations. Sample CVs were based on duplicate measurements of all available samples. Data were reported as median and interquartile range, unless stated otherwise.

| analyte | NfL |  | pNfH |  |
| --- | --- | --- | --- | --- |
| assay | homebrew Simoa | Quanterix Simoa | homebrew Simoa | Quanterix Simoa |
| analytical sensitivity | 2.4 pg/ml | 1.6 pg/ml | 3.2 pg/ml | 11.2 pg/ml |
| within-run precision | 7.4% (30.5 pg/ml)<br>7.6% (92.7 pg/ml)<br>6.2% (491.6 pg/ml) | 4.8% (10.1 pg/ml)<br>11.3% (51.6 pg/ml)<br>12.3% (155.1 pg/ml) | 2.9% (32.1 pg/ml)<br>4.9% (19.8 pg/ml)<br>6.2% (89.7 pg/ml) | 5.4% (18.5 pg/ml)<br>5.2% (267.1 pg/ml)<br>7.9% (1664 pg/ml) |
| between-run precision | 9.5% (30.5 pg/ml)<br>11.1% (92.7 pg/ml)<br>8.9% (491.6 pg/ml) | 7.2% (10.1 pg/ml)<br>11.3% (51.6 pg/ml)<br>15.4% (155.1 pg/ml) | 6.2% (32.1 pg/ml)<br>6.8% (19.8 pg/ml)<br>13.1% (89.7 pg/ml) | 12.1% (18.5 pg/ml)<br>7.5% (267.1 pg/ml)<br>9.9% (1664 pg/ml) |
| sample CV | 3.2% (1.1-5.4) | 4.1% (1.7-7.3) | 5.7% (3.1-11.0) | 4.2% (1.6-7.0) |

Assay validation in 47 independent serum samples (13 SCA3 subjects, 34 controls) demonstrated high agreement between the Quanterix and the homebrew NfL assays (R=0.99) (A). This allowed transformation of the NfL homebrew measurements to the scale of the Quanterix measurements by linear regression of log-transformed values and consecutive pooling of NfL values across the two cohorts. For pNfH, analogous validation showed lesser agreement between the Quanterix and the homebrew assays (R=0.88) (B), which we considered insufficient for transformation of pNfH homebrew measurements to the scale of the Quanterix measurements.

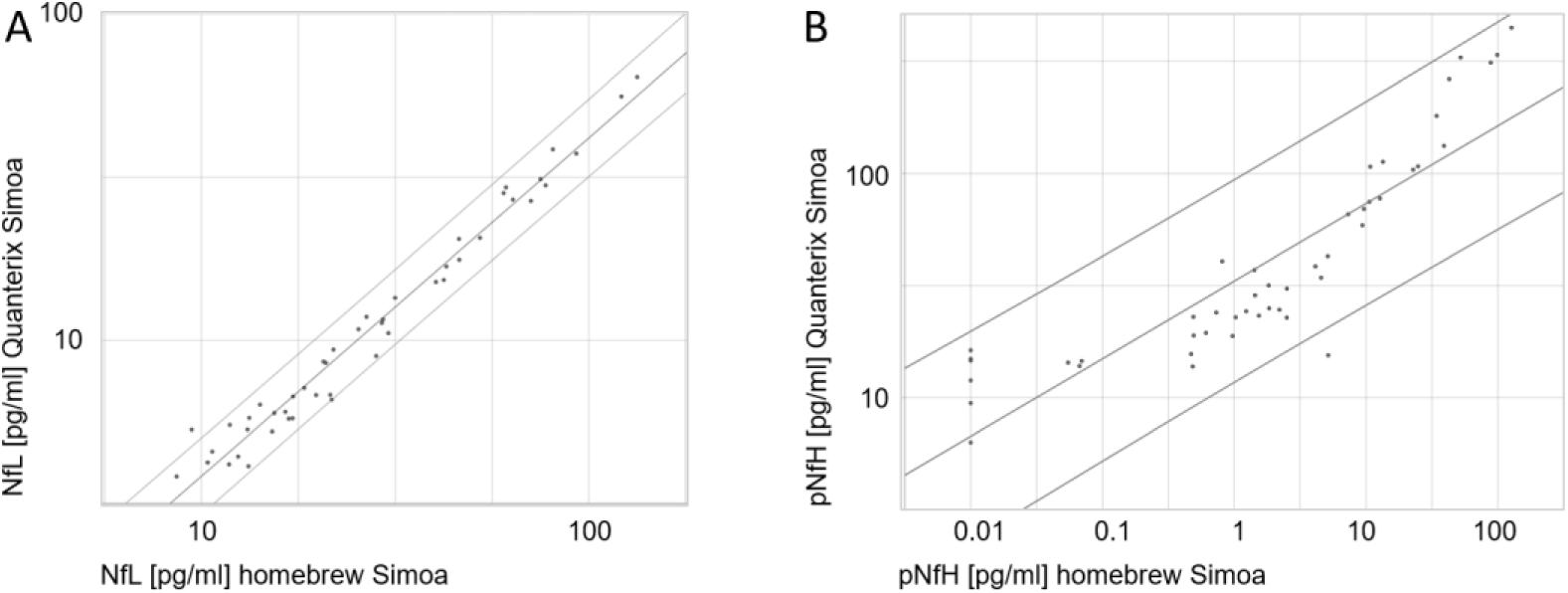

### Supplement 4. Sustained increase of NfL levels through the ataxic stage

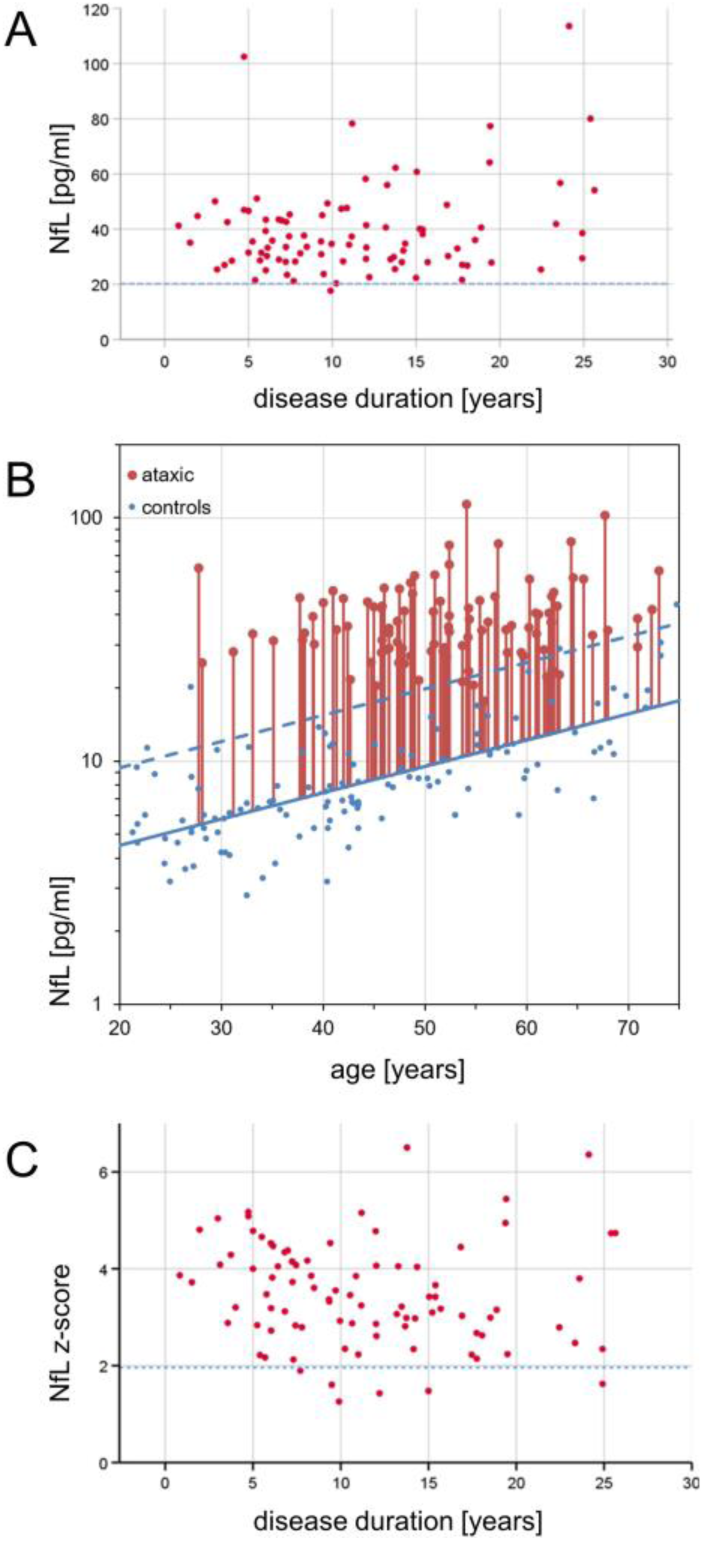

NfL levels of ataxic SCA3 subjects did not change significantly with disease duration (A), as indicated by linear regression using the pooled data of both cohorts (F(1,90)=1.56, p=.214, R^2^=0.02; slope: 1.37 ((−0.81)-3.55) (95% CI)) (A). This finding demonstrates a sustained increased NfL release from degenerating neurons throughout the disease course of SCA3. To compare ataxic SCA3 subjects with controls at the same age (B), we expressed the measured NfL level of SCA3 subjects as NfL z-score in relation to the age-dependent NfL distribution in controls. The NfL z-score of ataxic SCA3 subjects did not change significantly with disease duration (C), as revealed by linear regression (F(1,90)=0.18, p=.670, R^2^<0.01; slope: −0.01 ((−0.05)-0.03), indicating that NfL level remained increased through the ataxic stage of SCA3. As NfL levels increase with disease severity and disease progression rate (see main text), but not with disease duration (as shown here), they seem to capture the *functional* stage of SCA3 disease, rather than the mere calendrical estimate of duration of the disease. In addition, disease duration is an inherently difficult measure in ataxic patients as it relies on subjective, retrospective estimates.

### Supplement 5. Neurofilament plasma levels in heterozygous and homozygous SCA3 mice

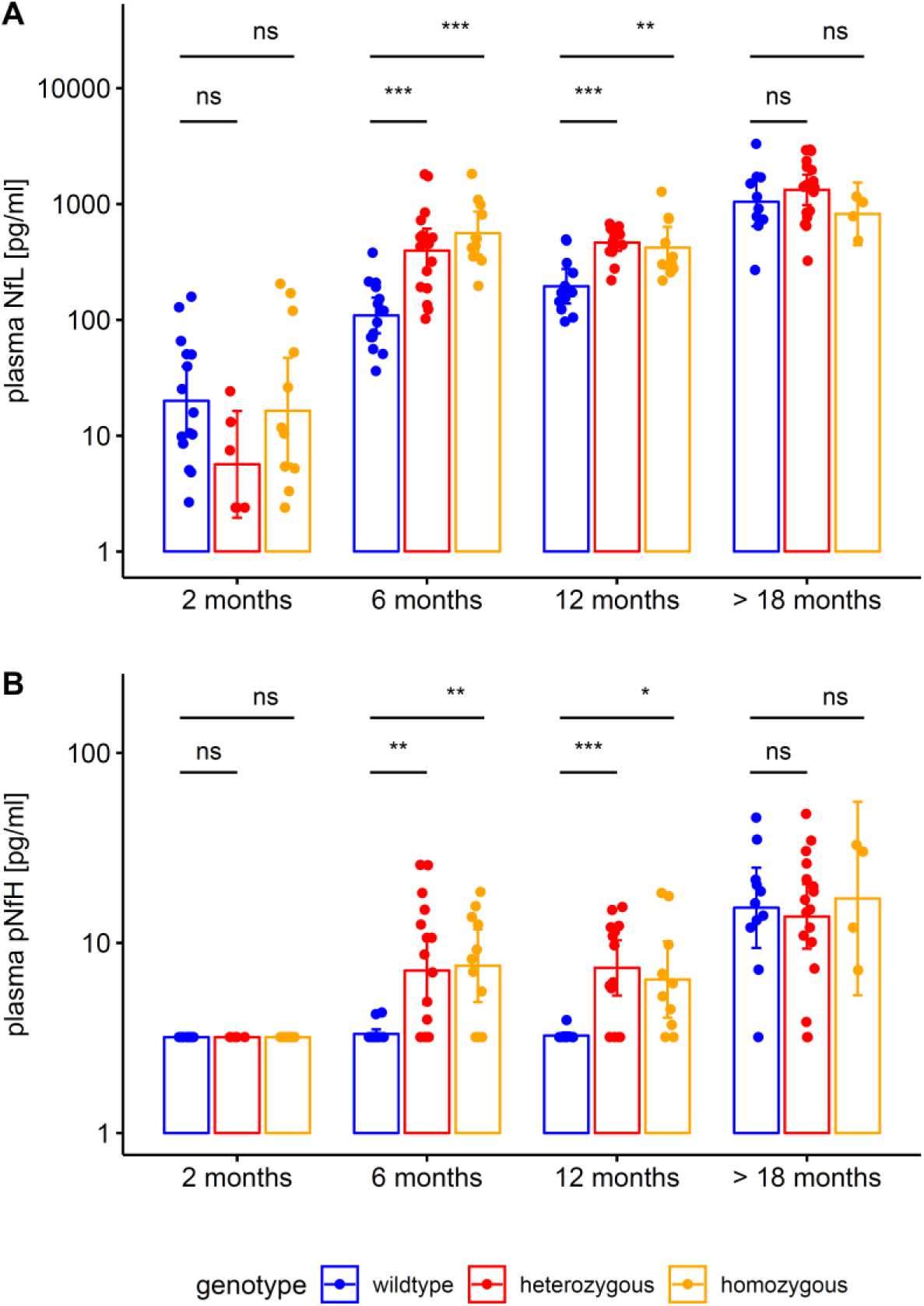

Plasma concentrations of NfL (A) and pNfH (B) (log-transformed, mean and 95% CI) were measured by Simoa assay in a SCA3 knock-in mouse model containing a 304-CAG-repeat in the *ATXN3* gene. Hetero- and homozygous animals were compared to wildtype animals by unpaired two-sample t tests, adjusted for unequal variances (*** p<.001, ** p<.01, ns p≥.05, Bonferroni-corrected). Homozygous mice confirmed the findings from heterozygous mice.

### Supplement 6. Brain tissue levels of neurofilaments in murine SCA3 disease

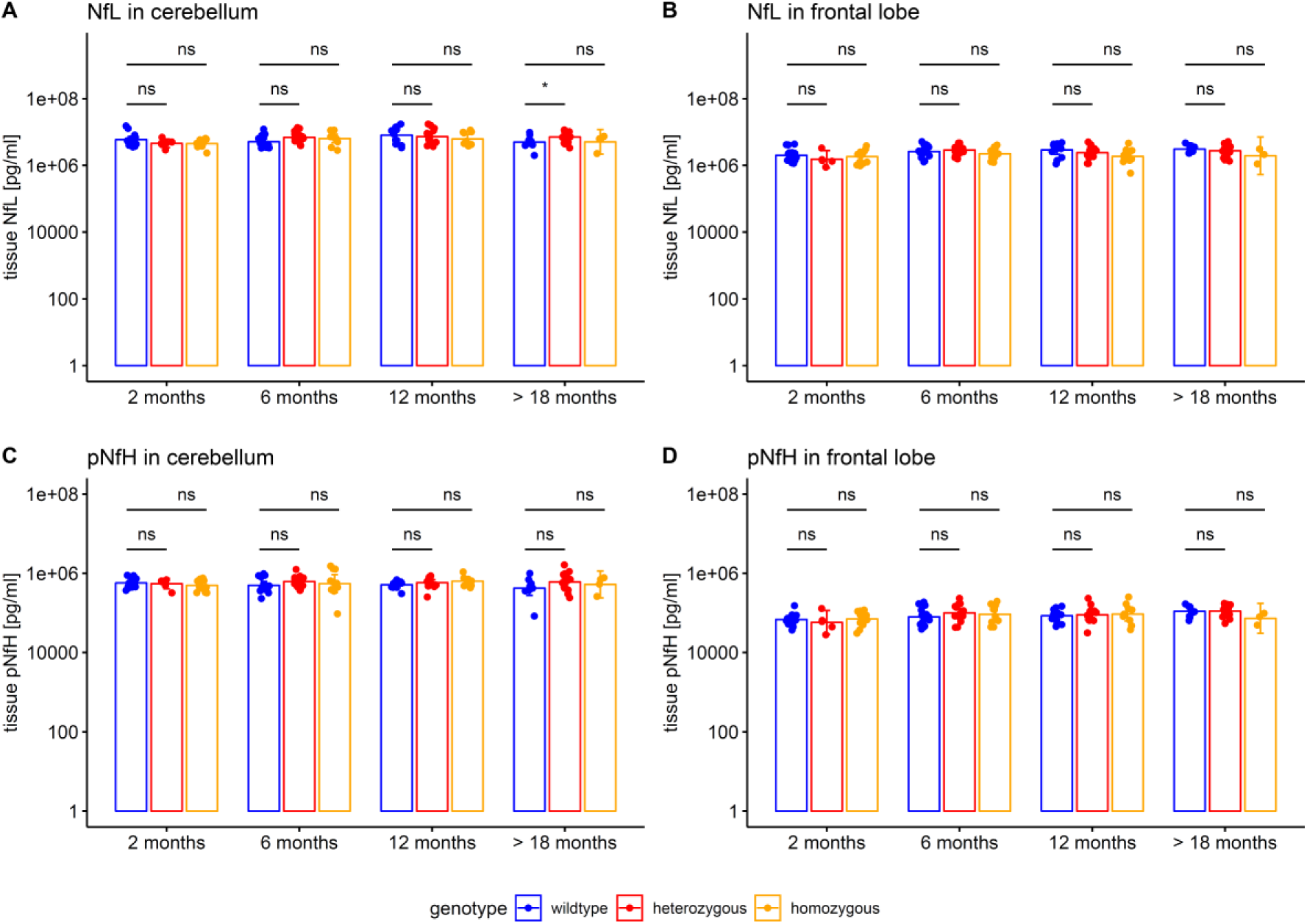

Neurofilament tissue concentrations (log-transformed, mean and 95% CI) in the cerebellum (A, C) and the frontal lobe (B, D) were measured by Simoa assay in the SCA3 mouse model. Hetero- and homozygous animals were compared to wildtype animals by unpaired two-sample t tests, adjusted for unequal variances (* p<.05, ns p≥.05, Bonferroni-corrected).

### Supplement 7. Weight phenotype in the 304Q knock-in SCA3 mouse model

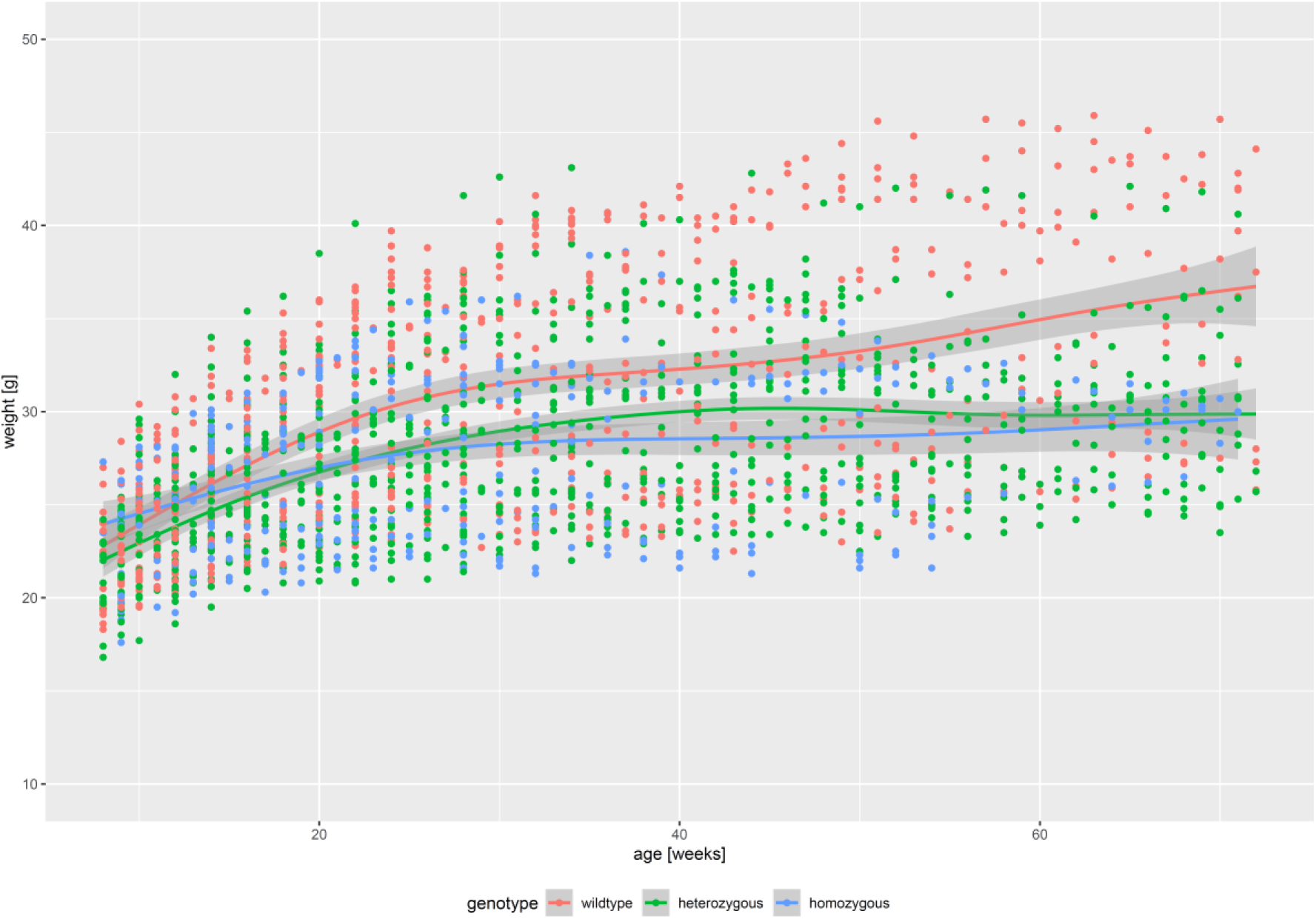

Weight was determined in all animals (n=145) every two weeks. For each genotype, weight was displayed as mean and 95% CI, as estimated by LOESS technique (locally estimated scatterplot smoothing, with standard span, as implemented in the R package ggplot2).

### Supplement 8. Motor phenotype in the 304Q knock-in SCA3 mouse model

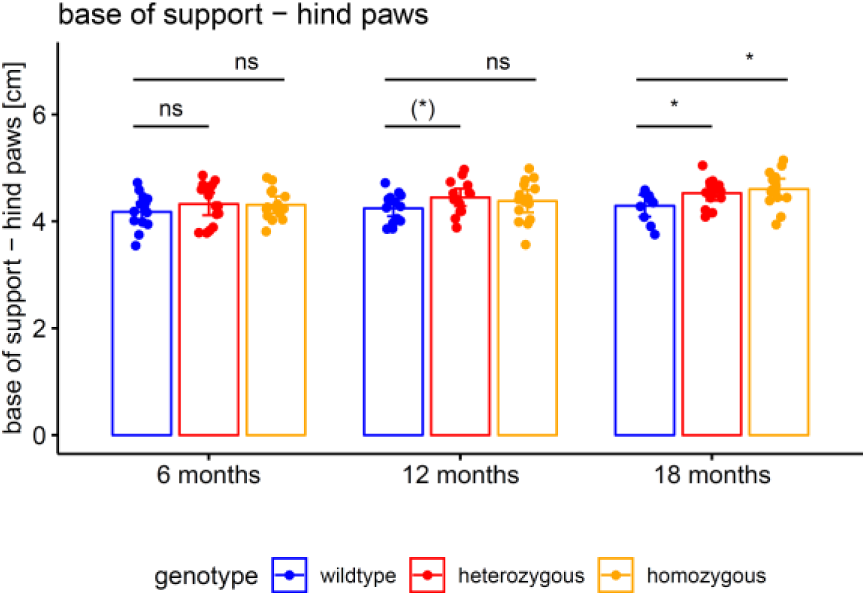

The motor phenotype of the 304Q knock-in SCA3 mouse model was assessed by automated gait analysis in the Catwalk system. Balance was captured by the base of support of the animal’s hind paws. Hetero- and homozygous animals were compared to wildtype animals by unpaired t tests (* p<.05, (*) p<.10, ns p≥.10).

### Supplement 9. Tissue levels of soluble and aggregated ataxin-3 in the cerebellum and frontal lobe in heterozygous and homozygous SCA3 mice

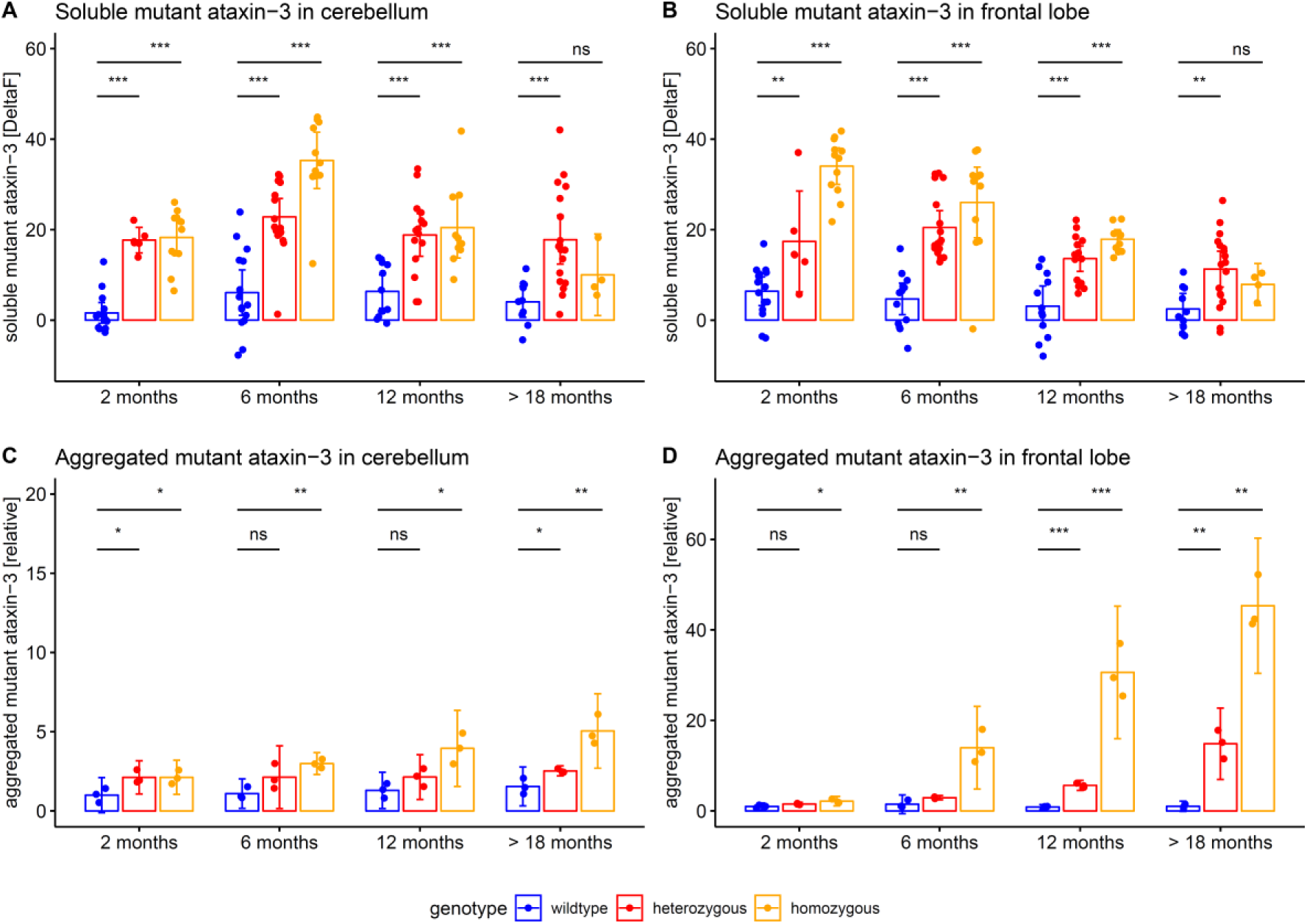

Tissue levels of soluble and aggregated mutant ataxin-3 (mean and 95% CI) were measured in cerebellum and frontal lobe. Hetero- and homozygous animals were compared to wildtype animals by unpaired t tests, adjusted for unequal variances (*** p<.001, ** p<.01, * p<.05, ns p≥.05). Homozygous mice confirmed the findings from heterozygous mice.

